# Monocyte Oxidative Stress Underlies Persistent Immune Activation in Long COVID Postural Orthostatic Tachycardia Syndrome

**DOI:** 10.64898/2026.05.08.26352776

**Authors:** Marwa A. Abd-Eldayem, Meenakshi Vinayagam, Yuliya A. Vance, Sachin Y. Paranjape, Celestine N Wanjalla, Kuniko C. Hunter, Sergey Dikalov, André Diedrich, Surat Kulapatana, Pouya E. Mehr, Tatiana X. Solis-Montenegro, David G. Harrison, Cyndya A. Shibao

## Abstract

Long COVID Postural Orthostatic Tachycardia Syndrome (LCPOTS) is characterized by persistent orthostatic tachycardia and systemic symptoms following SARS-CoV-2 infection. Many features of LCPOTS suggest ongoing immune activation, but the mechanisms driving this response remain unclear. In this study, we show that patients with LCPOTS, compared with individuals who recovered from SARS-CoV-2 without POTS, exhibit increased monocyte mitochondrial content and superoxide production, along with downregulation of NRF2-dependent antioxidant enzymes. This is accompanied by a marked increase in the formation of isolevuglandins (IsoLGs) in monocytes, which modify self-proteins and act as neoantigens capable of activating T cells. Consistent with this, LCPOTS patients exhibit a 3-fold increase in circulating T cell-monocyte doublets with immunological synapse formation. T cells in these complexes display a proinflammatory effector-memory and TEMRA phenotype, producing IFN-γ and IL-17A, which correlated with symptom severity. Circulating cytokines, including IL-17A, IFN-γ, and TNF-α, are elevated in patients with LCPOTS. In a subset of patients, transcutaneous vagal nerve stimulation (t-VNS) reduced circulating CD3^+^CD14^+^ doublets, IsoLG accumulation, and IL-6 expression in CD14^+^ monocytes. Our findings suggest that reduced vagal tone in LCPOTS leads to monocyte oxidative stress, IsoLG neoantigen formation, and T cell activation, linking immune dysregulation to cardiovagal dysfunction. Targeting these pathways may offer novel therapeutic opportunities.

**Graphical Abstract:** 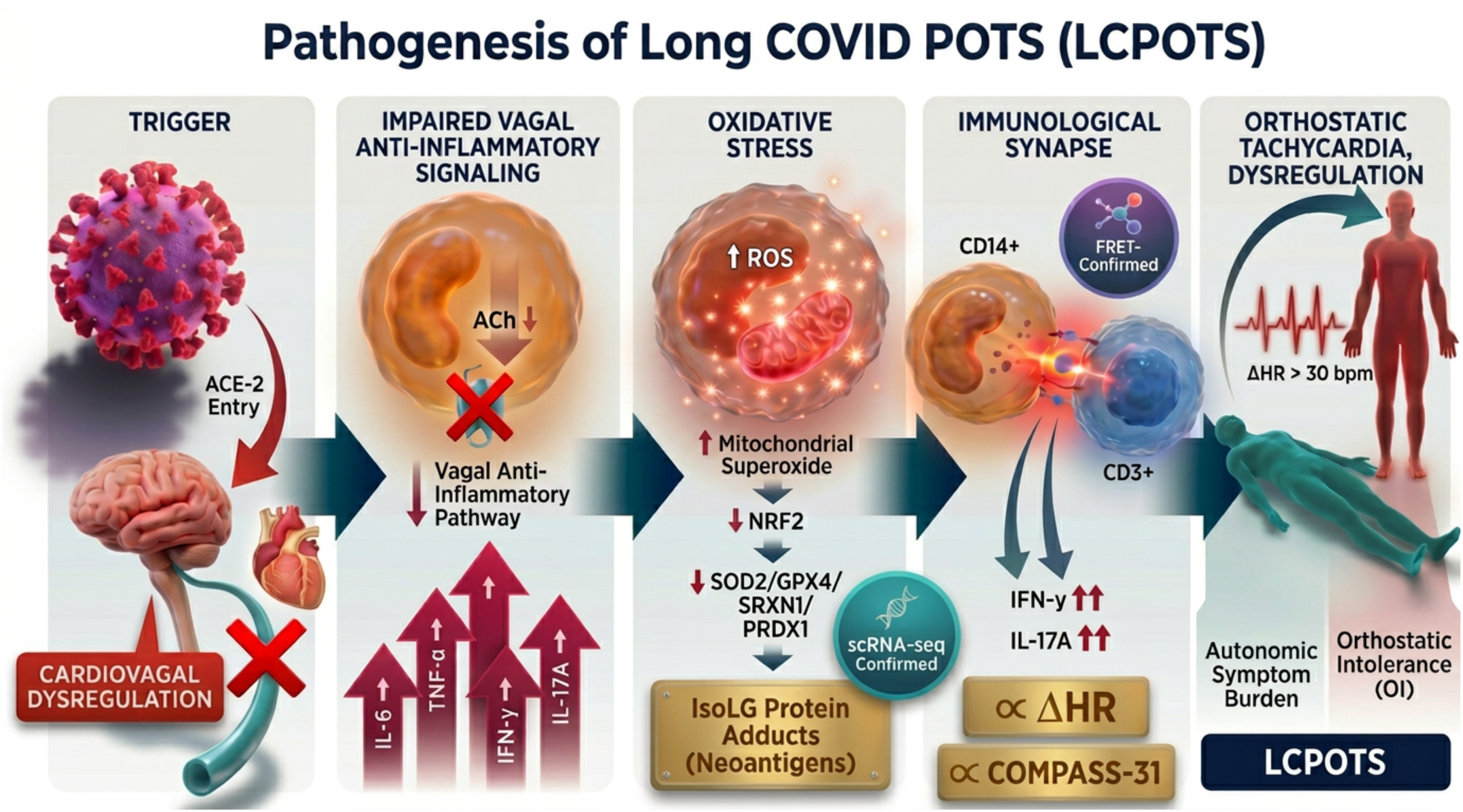

## Introduction

Postural orthostatic tachycardia syndrome (POTS) is a poorly understood condition that affects 1% of the population. It is associated with symptomatic tachycardia upon standing, and frequently with fatigue, chest pain, dyspnea, and gastrointestinal complaints (1, 2). POTS commonly occurs after viral infections, including those caused by the Epstein-Barr virus, gastrointestinal viruses, and influenza (3). The recent COVID-19 pandemic has led to a striking increase in POTS cases (4, 5), and it has been estimated that about 1/3^rd^ of patients with Long COVID-19 develops Long COVID POTS (LCPOTS) (6). About 80% of LCPOTS patients are female (7). This syndrome can persist for years and markedly impairs quality of life (8).

The causes of POTS and LCPOTS are poorly understood (9). One potential mechanism is viral infection of autonomic brain centers or nerves, leading to autonomic dysfunction even after viral clearance (10, 11). This is possible because autonomic centers possess angiotensin-converting enzyme-2 (ACE-2) receptors that allow entry of SARS-CoV-2 (12, 13). A second likelihood is that persistent immune activation and cytokine production can influence the function of both the sympathetic and parasympathetic nervous systems, leading to inappropriate vasodilatation, dysregulation of heart rate, and impaired blood volume control (14). In both LC and POTS, circulating cytokines, interleukin-6 (IL-6), tumor necrosis factor-α (TNF-α), and interferon-γ (IFN-γ), are elevated. Cytokines such as these can directly affect neural function. As an example, TNFα and IL-17A can induce demyelination in multiple sclerosis (15, 16) and viral infections (17), and IL-17A has been implicated in the autonomic dysfunction in Guillain-Barré syndrome (18). It is also possible that other products of activated immune cells, including reactive oxygen species (ROS), matrix metalloproteinases, and cytotoxic molecules, affect nerve function.

A potential mechanism for immune activation in LCPOTS is ongoing oxidative stress. Very recently, Shankar et al used several approaches to show that LC subjects and subjects with chronic fatigue syndrome have abnormal clearance pathways for reactive oxygen species (ROS) and that their higher ROS levels correlated with hyperproliferation of T cells and enhanced T cell function (19). Studies from our group have defined a novel mechanism by which ROS leads to immune activation. We have shown that excessive lipid peroxidation leads to the formation of highly reactive isolevuglandins (IsoLGs), which form stable protein adducts that act as neoantigens (20–22). These protein adducts are processed to IsoLG-adducted peptides that are presented in the context of both class 1 and class 2 major histocompatibility complexes to activate T cells. The accumulation of IsoLG-adducts in antigen-presenting cells is associated with the production of cytokines that skew T cells to produce both IL-17A and IFNγ (20). We have shown that these pathways contribute to the pathogenesis of hypertension (22), heart failure (21), and systemic lupus (23). The role of IsoLG-adducts in chronic conditions like LCPOTS remains undefined.

Related to the above, a recently identified marker of ongoing T cell activation is the presence of circulating T cell-monocyte doublets. These are increased in humans with active TB, in diabetic subjects with HIV, following vaccination, and in humans with Dengue fever (24, 25). Evidence suggests that these represent cells that have formed an immunological synapse, representing ongoing immune activation in these conditions. In the present study, we sought to determine if humans with LCPOTS, compared to those who had recovered from COVID without POTS, exhibit evidence of ongoing immune activation using this and other markers, and to determine if IsoLG-adducts contribute to this condition.

## Results

### Study Population, Baseline Characteristics, and Autonomic Symptom Burden Assessment

A total of 25 patients with LCPOTS and 15 subjects who recovered from COVID-19 without sequelae were studied. Demographic and baseline characteristics are summarized in Table 1. The groups were generally similar in age, body mass index, height, and weight. All participants in the LCPOTS group were female, whereas 80% of controls were female. Most participants in both groups were non-Hispanic and White. Baseline supine hemodynamic parameters were also similar between groups, with no significant differences in systolic or diastolic blood pressure or heart rate. Medications used by participants with LCPOTS included long-acting and short-acting β-blockers, midodrine, and guanfacine. These medications were stopped 2 weeks before study. Three LCPOTS patients also had hypertension (Table 1).

**Table 1.**
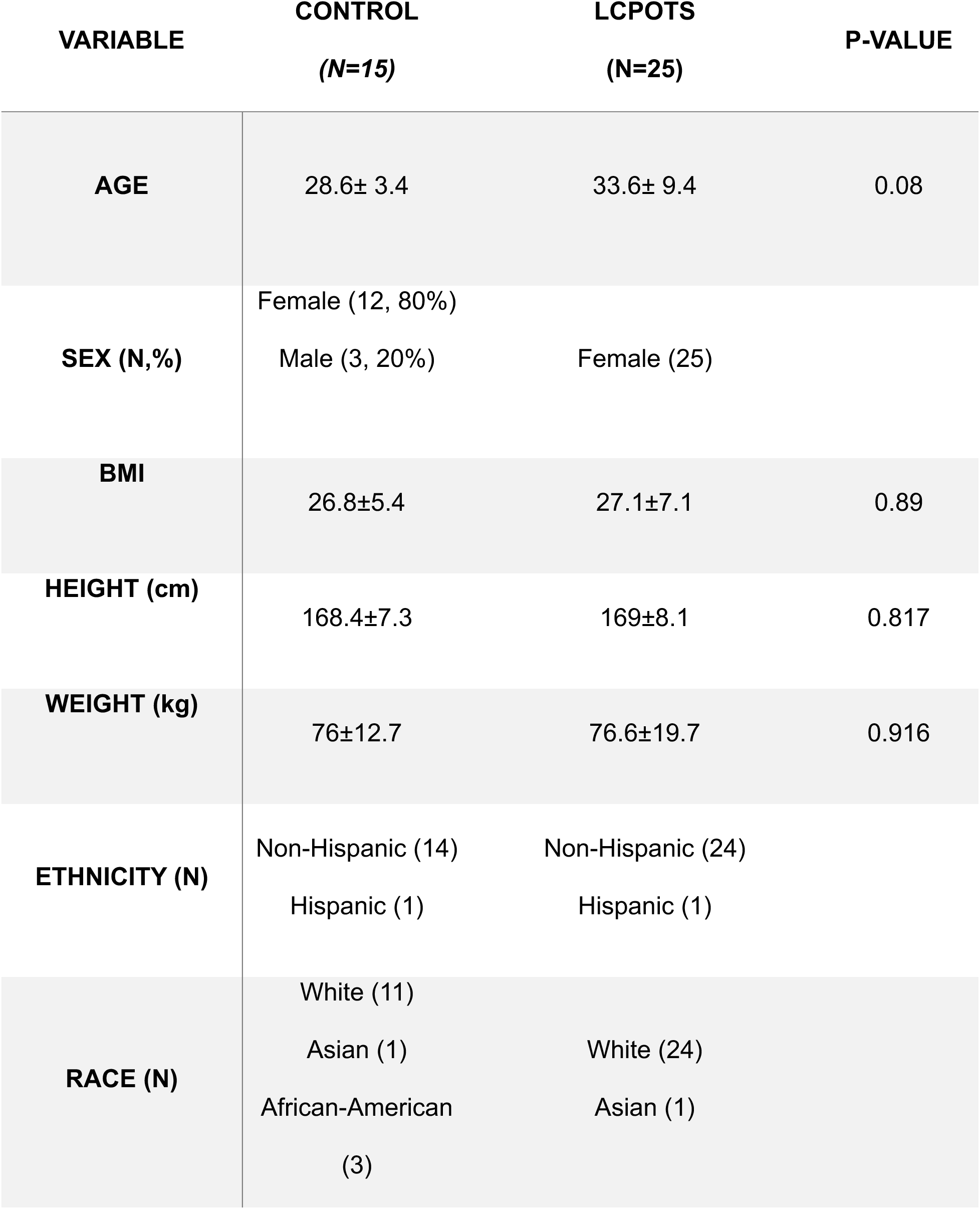
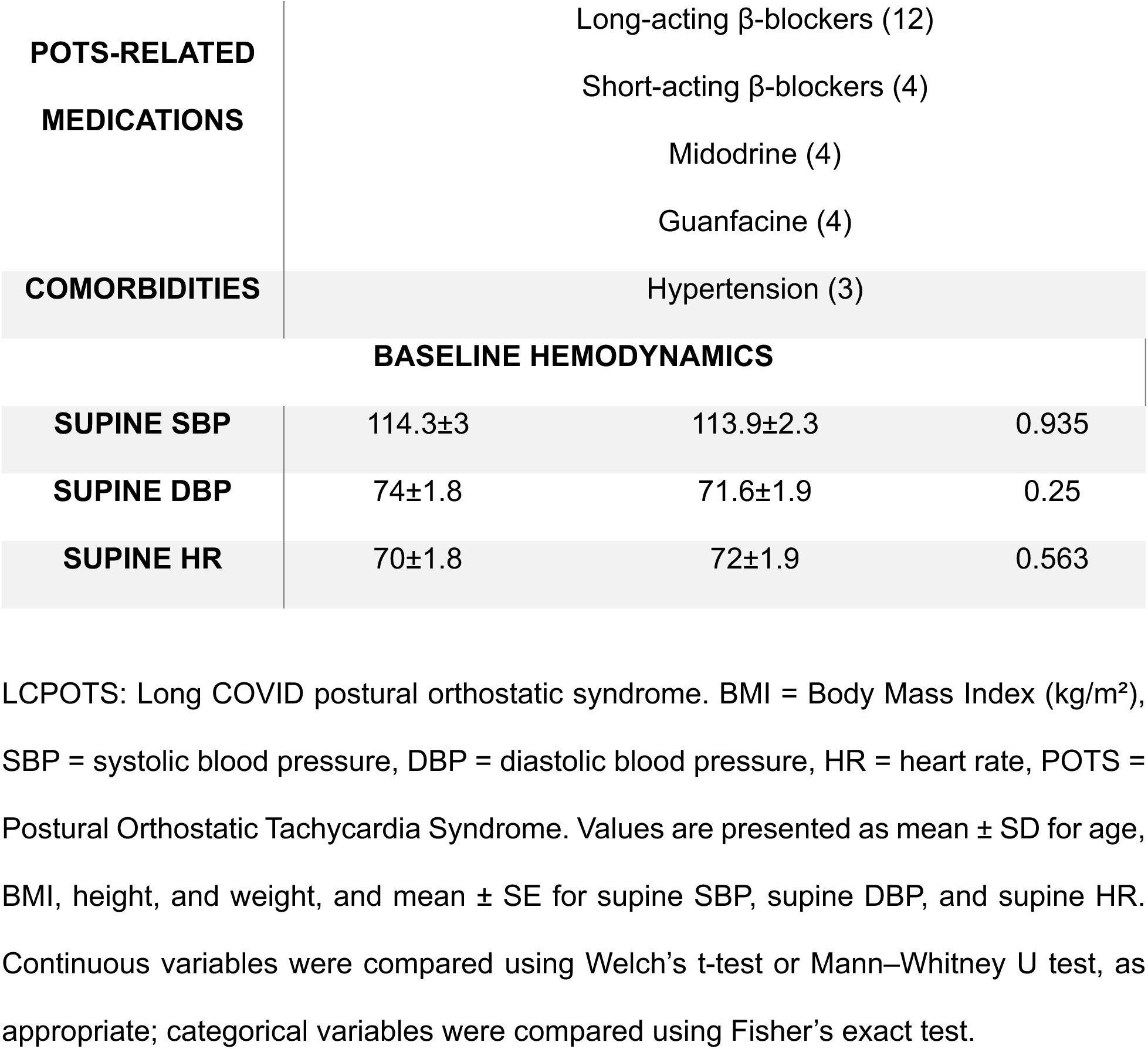
Demographic and Baseline Characteristics of Study Participants.

To define the clinical phenotype of LCPOTS, we examined orthostatic responses and orthostatic symptom burden. As shown in Figure 1, participants with LCPOTS exhibited greater orthostatic tachycardia and substantially higher autonomic symptom burden than recovered controls. Additional hemodynamic and symptom measures are shown in Figure 1S, A–F. COMPASS-31 scores were also markedly elevated in LCPOTS, with orthostatic intolerance representing the most prominent symptom domain, along with greater gastrointestinal, secretomotor, pupillomotor, vasomotor, and bladder symptom scores (Figure 1D and E).

**Figure 1.**
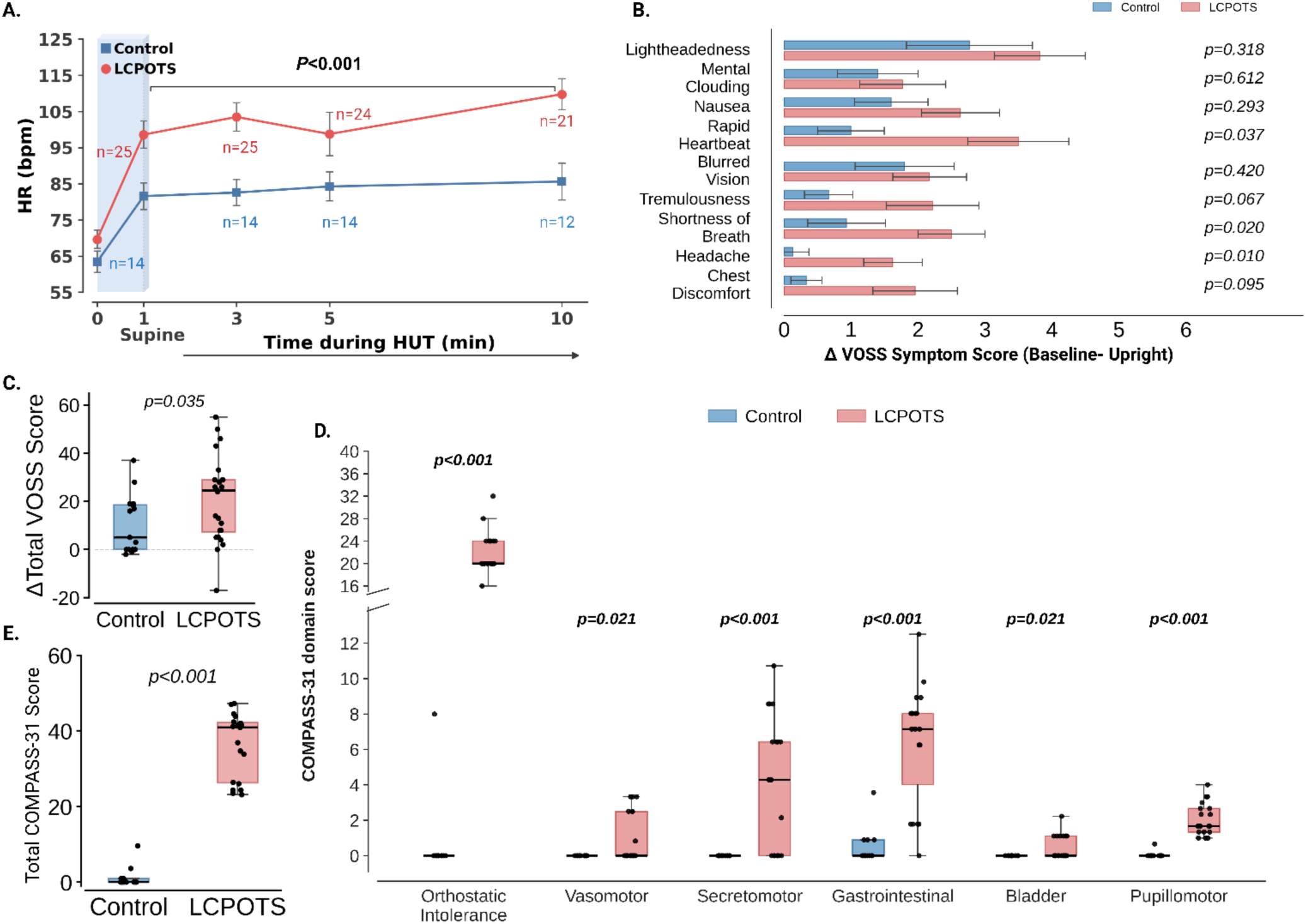
Comparative hemodynamic responses, orthostatic symptom burden, and autonomic symptom profile in healthy controls and participants with LCPOTS. **(A)** Heart rate (HR) during head-up tilt (HUT) in control and LCPOTS participants. Values are shown as mean ± SEM. The shaded region denotes the supine baseline period. Upright HR values (at 1, 3, 5, and 10 min) were analyzed using a Gaussian generalized estimating equation (GEE) model with exchangeable correlation structure, adjusted for time; the displayed *P* value represents the overall between-group effect across upright time points. Exact sample sizes at each time point are indicated on the graph. (B) Change in Vanderbilt Orthostatic Symptom Score (VOSS) symptom subscores from baseline to upright posture for individual symptom domains. Bars represent mean ± SEM. (C) Total change in VOSS score (ΔVOSS) in control and LCPOTS participants. (D) Composite Autonomic Symptom Score-31 (COMPASS-31) domain scores in controls and LCPOTS participants. (E) Total COMPASS-31 score in control and LCPOTS participants. COMPASS-31 domain and total scores were calculated using a validated method (29). For box-and-whisker plots (C–E), boxes indicate the interquartile range (25th–75th percentiles), center lines indicate the median, and whiskers extend to the most extreme values within 1.5 × the interquartile range; individual points represent individual participants. For panels B–E, between-group comparisons were performed using two-sided Mann–Whitney U tests; *P* values are shown on each panel. Sample sizes varied by measure because of symptom-data completeness (control *n*=12–15; LCPOTS *n*=17–24). HUT: head-up tilt; HR: heart rate; LCPOTS: Long COVID postural orthostatic tachycardia syndrome.

### Impaired Parasympathetic Modulation and Blunted Baroreflex Sensitivity in LCPOTS

LCPOTS was characterized by impaired cardiovagal modulation and reduced spontaneous baroreflex sensitivity. Although resting mean R-R interval and blood pressure were similar between groups (Figure 2A), participants with LCPOTS had significantly lower high-frequency power (HF) and total spectral power, whereas the LF/HF ratio did not differ significantly (Figure 2B). Time-domain measures, including RMSSD, SD1, and SD2, were also reduced, consistent with diminished cardiovagal modulation (Figure 2C). Blood pressure variability did not differ significantly between groups (Figure 2D). In contrast, baroreflex sensitivity for upward sequences was significantly attenuated, with a similar trend for downward sequences (Figure 2E).

**Figure 2:**
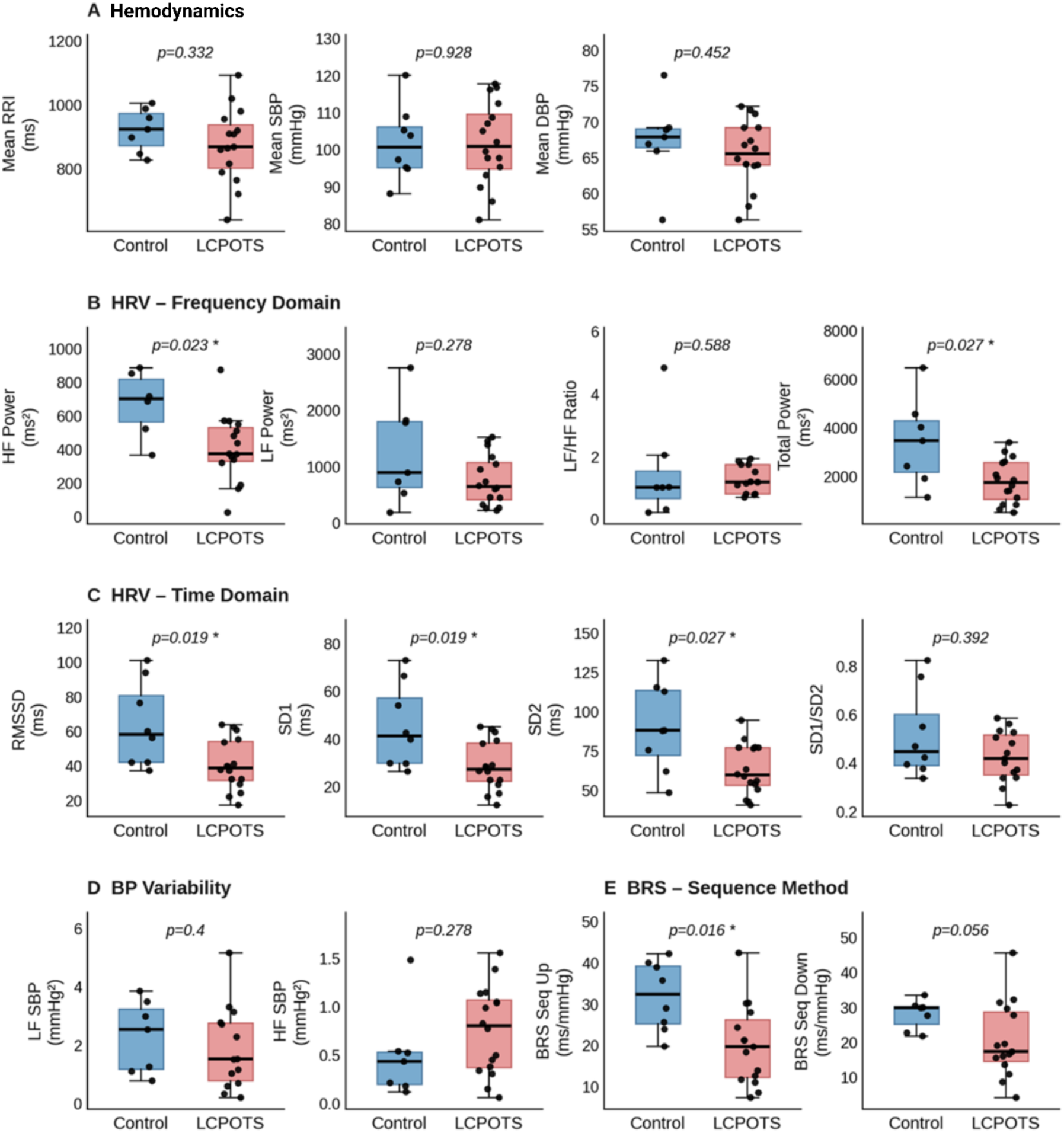
Autonomic cardiovascular profile in LCPOTS compared with healthy controls. Individual resting cardiovascular and autonomic measurements were available in a subset of healthy controls (blue, *n*=6-8) and participants with Long COVID postural orthostatic tachycardia syndrome (LCPOTS; red, *n*=13-16). **(A)** Hemodynamic parameters: mean R-R interval (RRI, ms), mean systolic blood pressure (SBP, mmHg), and mean diastolic blood pressure (DBP, mmHg). **(B)** Heart rate variability (HRV) frequency-domain measures: high-frequency power (HF Power, ms²), low-frequency power (LF Power, ms²), LF/HF ratio, and total spectral power (Total Power, ms²). **(C)** HRV time-domain measures: root mean square of successive differences (RMSSD, ms), Poincaré plot short-axis SD1 (ms), long-axis SD2 (ms), and SD1/SD2 ratio. **(D)** Blood pressure variability: LF component of systolic BP variability (LF SBP, mmHg²) and HF component (HF SBP, mmHg²). **(E)** Baroreflex sensitivity (BRS) estimated by the sequence method: upward sequences (BRS Seq Up, ms/mmHg) and downward sequences (BRS Seq Down, ms/mmHg). For box-and-whisker plots, boxes indicate the interquartile range (25th–75th percentiles), center lines indicate the median, and whiskers extend to the most extreme values within 1.5 × the interquartile range; individual points represent individual participants. Between-group comparisons were performed using two-sided Mann–Whitney U tests; *P* values are shown on each panel

### Preserved Valsalva Maneuver Hemodynamic Responses in LCPOTS

Autonomic reflex testing during the Valsalva maneuver is shown in Figure 1S, G–J. Controls had higher baseline systolic blood pressure during the maneuver, whereas baseline heart rate, phase II systolic blood pressure fall, late-phase II recovery, phase IV overshoot, and the Valsalva ratio were not significantly different between groups. Thus, despite clear evidence of orthostatic intolerance and impaired parasympathetic regulation in LCPOTS, standard Valsalva indices of sympathetic activity were largely preserved.

### scRNA-seq analysis of circulating leukocytes in LCPOTS

As an initial effort to understand if transcriptional programs are altered in circulating immune cells, we selected 4 female control subjects and 4 women with LCPOTS from our cohort. PBMCs from these subjects underwent scRNA-seq using the 10x Genomics platform. Principal component analysis and elbow-plot inspection identified approximately 9 major PBMC populations in both groups. Universal manifold approximation and projection plots demonstrated a broadly similar overall distribution of major immune cell populations and subpopulations in control subjects and women with LCPOTS (Figures 3A and 4S). Likewise, major differences in gene expression were not observed for T cells, NK cells, and B cells. In contrast, monocytes of LCPOTS subjects exhibited a striking alteration of gene expression (Figure 3B). Gene Ontology analysis of monocyte differentially expressed genes demonstrated enrichment for pathways related to leukocyte chemotaxis, cell migration, motility, and locomotion (Figure 3B-C), whereas pathway analysis highlighted antigen processing and presentation, lysosomal/phagosomal pathways, and other immune-related programs (Figure 3D). Before quality-control filtering, PBMCs from LCPOTS showed increased mitochondrial transcript content compared with controls (Figure 3E, upper panel). This difference between Controls and LCPOTS persisted after exclusion of cells with >10% mitochondrial transcripts (Figure 3E, lower panel). Differential expression analysis revealed decreased expression of multiple antioxidant and redox-defense genes in CD14^+^ monocytes from women with LCPOTS, including NFE2L2, SOD2, GPX4, PRDX1, and HMOX1 (Figure 3F–J and Table 2S). Additional antioxidant-related genes were also downregulated (Table 2S).

**Figure 3.**
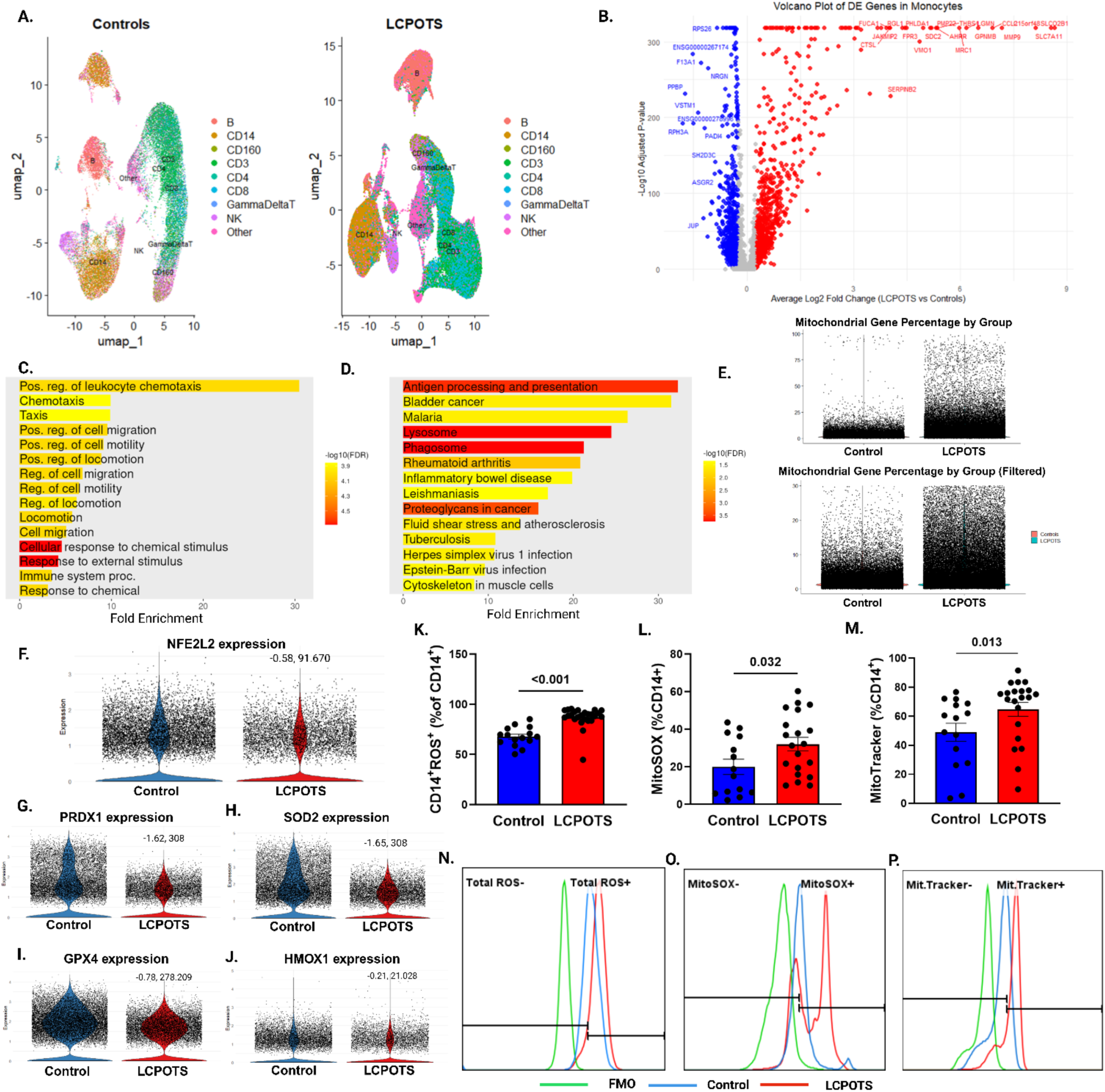
Monocyte transcriptional remodeling and increased oxidative stress in LCPOTS. **(A)** Uniform manifold approximation and projection (UMAP) visualization of peripheral blood mononuclear cells (PBMCs) from control participants (left) and patients with long COVID postural orthostatic tachycardia syndrome (LCPOTS, right), colored by major immune cell clusters (B cells, CD14^+^ monocytes, CD160^+^ cells, CD3^+^ T cells, CD4^+^ T cells, CD8^+^ T cells, γδ T cells, NK cells, and Other). **(B)** Volcano plot of differentially expressed genes (DE) in monocytes from LCPOTS versus controls; red indicates genes upregulated in LCPOTS, and blue indicates genes downregulated in LCPOTS. Selected top-ranked DE genes are labeled. The x-axis shows the average log_2_ fold change (LCPOTS vs. controls) and the y-axis shows the −log_10_ adjusted *P* value. **(C)** Gene Ontology (GO) biological process enrichment analysis of monocyte DE genes, showing enrichment of processes related to leukocyte chemotaxis, cell migration, motility, and locomotion. Bar color indicates −log_10_ false discovery rate (FDR). **(D)** Pathway enrichment analysis (KEGG) of monocyte DE genes, showing enrichment of antigen processing and presentation, lysosome and phagosome pathways, together with broader immune and inflammation-related pathways. Bar color indicates −log_10_ FDR. **(E)** Quality-control assessment showing the percentage of mitochondrial transcripts per cell in controls and LCPOTS, before (upper panel) and after (lower panel) filtering of low-quality cells. Each dot represents one cell. **(F–J)** Violin plots showing expression of **NFE2L2** (**F**), **PRDX1** (**G**), **SOD2** (**H**), **GPX4** (**I**), and **HMOX1** (**J**) in CD14^+^ monocytes from controls (Blue) and LCPOTS (Red). Values shown in each panel denote the average log_2_ fold change (LCPOTS vs. controls) and the corresponding log_10_ adjusted *P* value from the differential expression analysis. Single-cell RNA sequencing(scRNA-seq) was performed on PBMCs from 4 female control subjects and 4 women with LCPOTS. Differential gene expression was analyzed using the Wilcoxon rank-sum test with Bonferroni correction for multiple testing. Pathway and GO enrichment analyses were corrected using the Benjamini–Hochberg false discovery rate. **(K–M)** Flow cytometric quantification of total intracellular reactive oxygen species (ROS)-positive **(K)**, mitochondrial superoxide (MitoSOX)-positive **(L)**, and MitoTracker-positive **(M)** CD14^+^ monocytes, expressed as a percentage of CD14^+^ cells. Bars represent mean ± SEM, and individual dots represent individual participants. Between-group comparisons were performed using two-sided Mann–Whitney U tests; *P* values are shown above each comparison. **(N–P)** Representative flow cytometry histograms of total ROS **(N)**, MitoSOX **(O)**, and MitoTracker **(P)** in CD14^+^ monocytes from controls (blue) and LCPOTS (red), with fluorescence-minus-one (FMO) controls shown in green. Vertical gates were used to define positive populations. scRNA-seq, *n* = 4 controls and *n* = 4 LCPOTS (all female); flow cytometry, controls *n*=14-15, LCPOTS *n* = 20–25 depending on the availability of samples.

**Figure 4:**
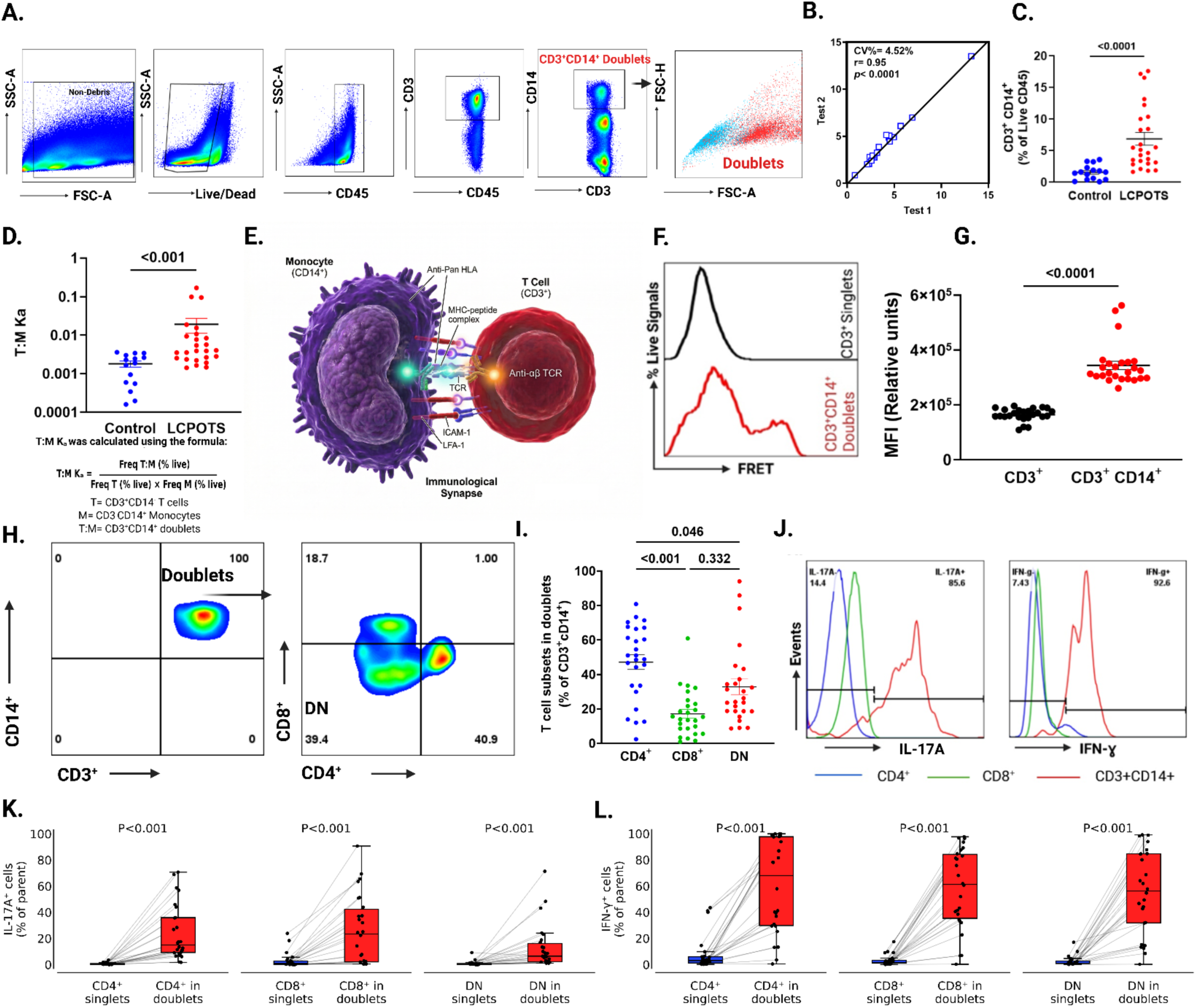
Circulating T cell-monocyte doublets as markers of persistent inflammation in LCPOTS. Individual flow cytometric analyses were performed in controls and participants with long COVID postural orthostatic tachycardia syndrome (LCPOTS). **(A)** Representative gating strategy showing sequential identification of non-debris, live CD45^+^ leukocytes, discrimination of singlets and doublets, and definition of CD3^+^CD14^+^ population within the doublet gate. **(B)** Reproducibility of CD3^+^CD14^+^ doublet measurements in two consecutive assays from 17 subjects. Doublets, expressed as a percentage of live CD45^+^ cells, showed low variability and strong reproducibility between measurements (Spearman correlation). **(C)** Frequency of circulating doublets (CD3^+^CD14^+^) expressed as a percentage of live CD45^+^ cells in the PBMCs of LCPOTS patients compared to controls. **(D)** T cell-monocyte association constant (T:M K_a_) in controls and LCPOTS participants. T:M Kₐ was calculated as the frequency of CD3^+^CD14^+^ doublets divided by the product of the frequencies of CD3^+^CD14^-^ T cells and CD3^-^CD14^+^ monocytes among live CD45^+^ cells (25). **(E)** Schematic illustration of fluorescence resonance energy transfer (FRET)-based detection of immunological synapse formation between T cells and monocytes in CD3^+^CD14^+^ doublets using anti-α/β TCR and anti-pan HLA conjugated to a complementary FRET fluorophore pair. FRET signal detection in CD3^+^CD14^+^ doublets was compared with that in CD3^+^ cells alone, which served as a negative control. **(F)** Representative FRET histograms showing minimal signal in CD3^+^ singlets and increased signal in CD3^+^CD14^+^ doublets. **(G)** TCR-HLA FRET proximity mean fluorescence intensity (MFI) in CD3^+^ singlets and CD3^+^CD14^+^ doublets. **(H)** Representative gating strategy for identification of CD3^+^CD14^+^ doublets (left) and subdivision of the doublet population into CD4^+^, CD8^+^, and double-negative (DN) T cell subsets (right). **(I)** Quantification of CD4^+^, CD8^+^, and DN T cell subsets within CD3^+^CD14^+^ doublets, expressed as a percentage of total CD3^+^CD14^+^ cells. **(J)** Representative intracellular cytokine staining histograms showing IL-17A (left) and IFN-γ (right) in CD4^+^ (blue), CD8^+^ (green), and CD3^+^CD14^+^ (red) populations. **(K and L)** Paired comparisons of IL-17A^+^ (**K**) and IFN-γ^+^ (**L**) cells among CD4^+^, CD8^+^, and DN singlet T cells versus their corresponding populations within CD3^+^CD14^+^ doublets. Individual lines connect matched singlet–doublet pairs from the same participant. For panels (C) and (D), between-group comparisons were performed using two-sided Mann– Whitney U tests. For panel (F), comparisons were performed using a two-sided Wilcoxon signed-rank test. For panel (I), comparisons across CD4^+^, CD8^+^, and DN subsets were performed using the Friedman test followed by Dunn’s multiple-comparisons correction. For panels (K) and (L), matched singlet versus doublet comparisons within each T cell subset were analyzed using two-sided Wilcoxon signed-rank tests, with Holm correction for multiple comparisons across CD4^+^, CD8^+^, and DN subsets. *P* values are shown on the corresponding panels. Controls, n = 15; LCPOTS, n = 25, unless otherwise indicated.

**Table 2:** Plasma cytokine profiles in LCPOTS patients and healthy controls.

| Cytokine | Control | LCPOTS | P-value |
| --- | --- | --- | --- |
| **IL-17A** | 0.189 [0.135–0.355] | 0.677 [0.445–0.915] | 0.001 |
| **IL-6** | 0.609 [0.473–0.887] | 1.31 [0.884–1.57] | 0.001 |
| **TNF-α** | 0.390 [0.334–0.484] | 0.509 [0.452–0.676] | 0.009 |
| **IL-12/IL-23p40** | 66.3 [48.0–93.8] | 95.9 [81.1–164.9] | 0.018 |
| **IFN-γ** | 6.23 [5.05–8.05] | 9.01 [6.68–11.9] | 0.025 |
| **IL-8** | 5.50 [4.53–5.91] | 6.11 [5.28–8.08] | 0.043 |
| **IL-1β** | 0.135 [0.087–0.196] | 0.216 [0.155–0.333] | 0.044 |
| **IL-10** | 0.176 [0.135–0.223] | 0.218 [0.164–0.313] | 0.104 |
| **IL-4** | 0.013 [0.005–0.041] | 0.017 [0.011–0.024] | 0.988 |

LCPOTS: Long COVID postural orthostatic syndrome. controls n=8–12; LCPOTS n=16–18. Values are presented as median [Q1, Q3] expressed in pg/mL, Mann-Whitney test

### Production of reactive oxygen species (ROS) by monocytes of LCPOTS subjects

Given the striking increase in mitochondrial number and the marked reduction in antioxidant gene expression identified by scRNA sequencing, we next quantified reactive oxygen species (ROS) in monocytes from LCPOTS subjects (Figure 3). Total intracellular ROS and mitochondrial superoxide were measured by flow cytometry using the redox-sensitive probes CellROX and MitoSOX, respectively. As summarized in the mean data (Figure 3K) and representative histograms (Figure 3N), total ROS levels were significantly elevated in monocytes from LCPOTS subjects compared with controls. Mitochondrial superoxide production was also approximately twofold higher in LCPOTS monocytes (Figure 3L and 3O). Notably, in keeping with the expanded mitochondrial transcript content observed in the 10x scRNA-seq analysis, mitochondrial content was significantly increased by the MitoTracker assay for all our subjects (Figure 3M and P).

### T Cell-Monocyte Doublets Are Increased in LCPOTS and Form Functional Immune Synapses

Circulating T cell-monocyte doublets were identified as the CD3^+^CD14^+^ double-positive population using the gating strategy shown in Figure 4A. Repeated measurements demonstrated high reproducibility of doublet quantification (Figure 4B). Using this approach, we observed a significant increase in circulating CD3^+^CD14^+^ doublets in patients with LCPOTS compared with controls (Figure 4C). In parallel, the T cell-monocyte association constant (T:M Kₐ) was significantly higher in LCPOTS than in controls (Figure 4D), indicating enhanced association between circulating T cells and monocytes in LCPOTS. T:M K_a_ was calculated as indicated beneath Figure 4D. The FRET strategy used to detect immunological synapse formation is illustrated in Figure 4E. CD3^+^CD14^+^ doublets exhibited significantly higher TCR-HLA FRET mean fluorescence intensity (MFI) than CD3^+^ singlets (Figure 4F), and representative histograms showed a clear FRET MFI signal in doublets compared to CD3^+^ singlets, which serve as a negative control (Figure 4G). Together, these findings indicate that the expanded CD3^+^CD14^+^ population in LCPOTS represents true T cell-monocyte immune synapses rather than random cellular aggregates.

### T Cells Within Doublets Display a Proinflammatory Phenotype

CD3^+^CD14^+^ doublets contained CD4^+^, CD8^+^, and double-negative (DN) T cell populations, as shown by the gating strategy in Figure 4H and quantified in Figure 4I, indicating that multiple T cell subsets participate in monocyte interactions in LCPOTS. Among subjects with LCPOTS, intracellular cytokine staining showed that T cells engaged in CD3^+^CD14^+^ doublets exhibited a markedly more proinflammatory phenotype than their corresponding singlet populations. Representative histograms demonstrated rightward shifts in intracellular IL-17A and IFN-γ staining in doublet-associated T cells relative to singlet T cell populations (Figure 4J). The frequency of IL-17A^+^ cells was significantly higher in doublet than singlet T cell subsets, including CD4^+^, CD8^+^, and DN T cells (Figure 4K). Similarly, the frequency of IFN-γ^+^ cells was significantly higher in doublet-associated than in singlet CD4^+^, CD8^+^, and DN T cell subsets (Figure 4L). Together, these data indicate that T cells within CD3^+^CD14^+^ doublets are enriched for inflammatory cytokine production, consistent with enhanced activation in the context of T cell–monocyte interactions.

### Cytokine-Positive Doublets Correlate with Orthostatic Tachycardia and Autonomic Symptom Burden

We next examined whether this inflammatory phenotype was related to clinical severity. The frequency of both CD3^+^CD14^+^IL-17A^+^ and CD3^+^CD14^+^IFN-γ^+^ doublets correlated positively with the magnitude of maximum heart rate increase during orthostatic challenge (Figure 5A and B). Similarly, the frequencies of CD3^+^CD14^+^IL-17A^+^ and CD3^+^CD14^+^IFN-γ^+^ doublets correlated positively with total COMPASS-31 score (Figure 5C and D). These data suggest that proinflammatory activation of T cells within CD3^+^CD14^+^ doublets is linked to orthostatic tachycardia and greater autonomic symptom burden in LCPOTS.

**Figure 5:**
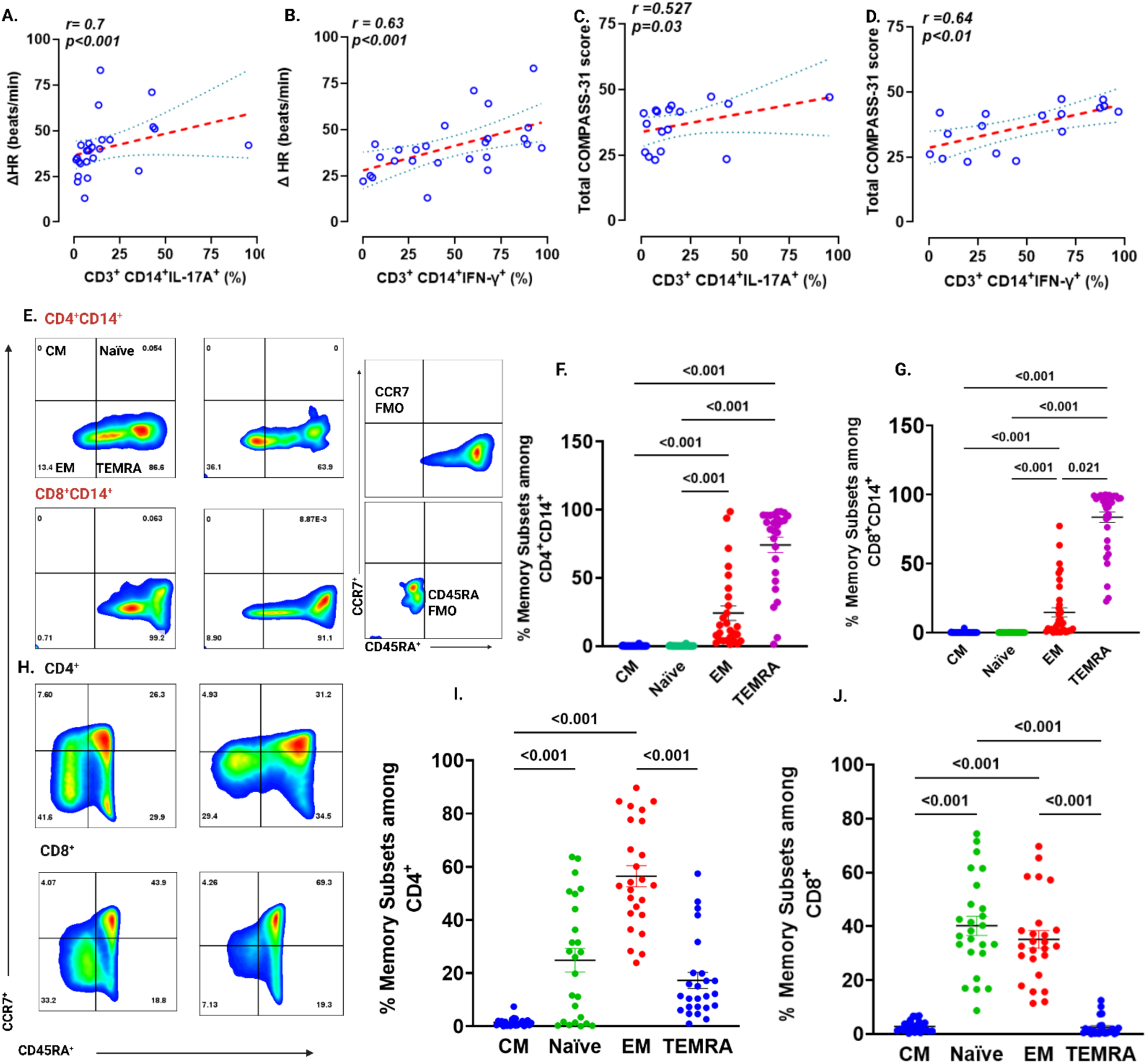
Cytokine-positive CD3^+^CD14^+^ doublets are associated with orthostatic tachycardia and autonomic symptom burden and are enriched for EM/TEMRA phenotypes in LCPOTS. Analyses were performed in participants with long COVID postural orthostatic tachycardia syndrome (LCPOTS). (**A and B)** Correlation of ΔHR (beats/min) with the percentages of CD3^+^CD14^+^IL-17A^+^ cells and CD3^+^CD14^+^IFN-γ^+^ cells, respectively (n=24). (**C and D)** Correlations of total COMPASS-31 score with the percentages of CD3^+^CD14^+^IL-17A^+^ cells and CD3^+^CD14^+^IFN-γ^+^ cells, respectively (n=17). For Panels A - D, associations were assessed with 2-sided Spearman rank correlation; corresponding *r* and *P* values are shown in each panel. Red dashed lines indicate fitted regression lines, and blue dotted lines indicate 95% CIs. (**E**) Representative flow cytometry plots showing memory-subset distribution within CD4^+^CD14^+^ cells (top) and CD8^+^CD14^+^ cells (bottom) from 2 different participants, with fluorescence-minus-one (FMO) controls for CCR7 and CD45RA shown at right. Subsets were defined as central memory (CM; CCR7^+^CD45RA^-^), naïve (CCR7^+^CD45RA^+^), effector memory (EM; CCR7^-^CD45RA^-^), and terminally differentiated effector memory re-expressing CD45RA (TEMRA; CCR7^-^CD45RA^+^). **(F)** Quantification of memory subsets among CD4^+^CD14^+^ cells. (**G**) Quantification of memory subsets among CD8^+^CD14^+^ cells. **(H)** Representative flow cytometry plots showing memory-subset distribution within singlets CD4^+^ cells (top) and CD8^+^ cells (bottom) from 2 different participants. **(I)** Quantification of memory subsets among CD4^+^ cells. **(J)** Quantification of memory subsets among CD8^+^ cells. For **F, G, I, and J**, data are mean ± SEM; statistical comparisons were performed using a Friedman test followed by Dunn multiple-comparisons correction. Exact *P* values are shown above the relevant comparisons (n = 25).

### T Cell Memory Phenotypes Within CD3^+^CD14^+^ Doublets Are Skewed Toward EM/TEMRA Subsets

To characterize T cell phenotypes within doublet populations, we analyzed 2 key surface markers: CD45RA, which distinguishes naïve from memory T cell populations, and CCR7, a lymphoid-homing receptor that defines memory T cell subsets. Representative memory-phenotyping plots from two different patients, together with fluorescence-minus-one controls for CCR7 and CD45RA, are shown in Figure 5E. Quantification demonstrated that both CD4^+^CD14^+^ and CD8^+^CD14^+^ doublets were predominantly composed of effector memory (EM; CD45RA^-^CCR7^-^) and terminally differentiated effector memory re-expressing CD45RA (TEMRA; CD45RA^+^CCR7^-^) cells, with relatively few central memory (CM; CD45RA^-^CCR7^+^) or naïve (CD45RA^+^CCR7^+^) cells (Figure 5F and G). Overall, EM and TEMRA cells accounted for approximately 90% of T cells within doublets. In contrast, TEMRA-like cells were much less common in the singlet populations from the same subjects (Figure 5 H-J). This bias toward TEMRA cells suggests that these cells have undergone multiple rounds of activation and proliferation, likely due to repeated antigen exposure.

### T cell-monocyte doublets and IsoLG-adduct accumulation

Because we observed increased ROS formation in monocytes of subjects with LCPOTS, we considered the hypothesis that this leads to IsoLG formation, which in turn forms adducts with self-proteins and neoantigen formation. As shown in the example histogram (Figure 6A) and mean data (Figure 6B), IsoLG-adducts were markedly higher in T cell-monocyte doublets compared to monocyte singlets or in T cells alone. Analysis of the three monocyte subsets demonstrated a higher percentage of IsoLG-adduct-positive cells in classical, intermediate, and non-classical monocytes from LCPOTS patients compared to controls (Figure 6C-E).

**Figure 6:**
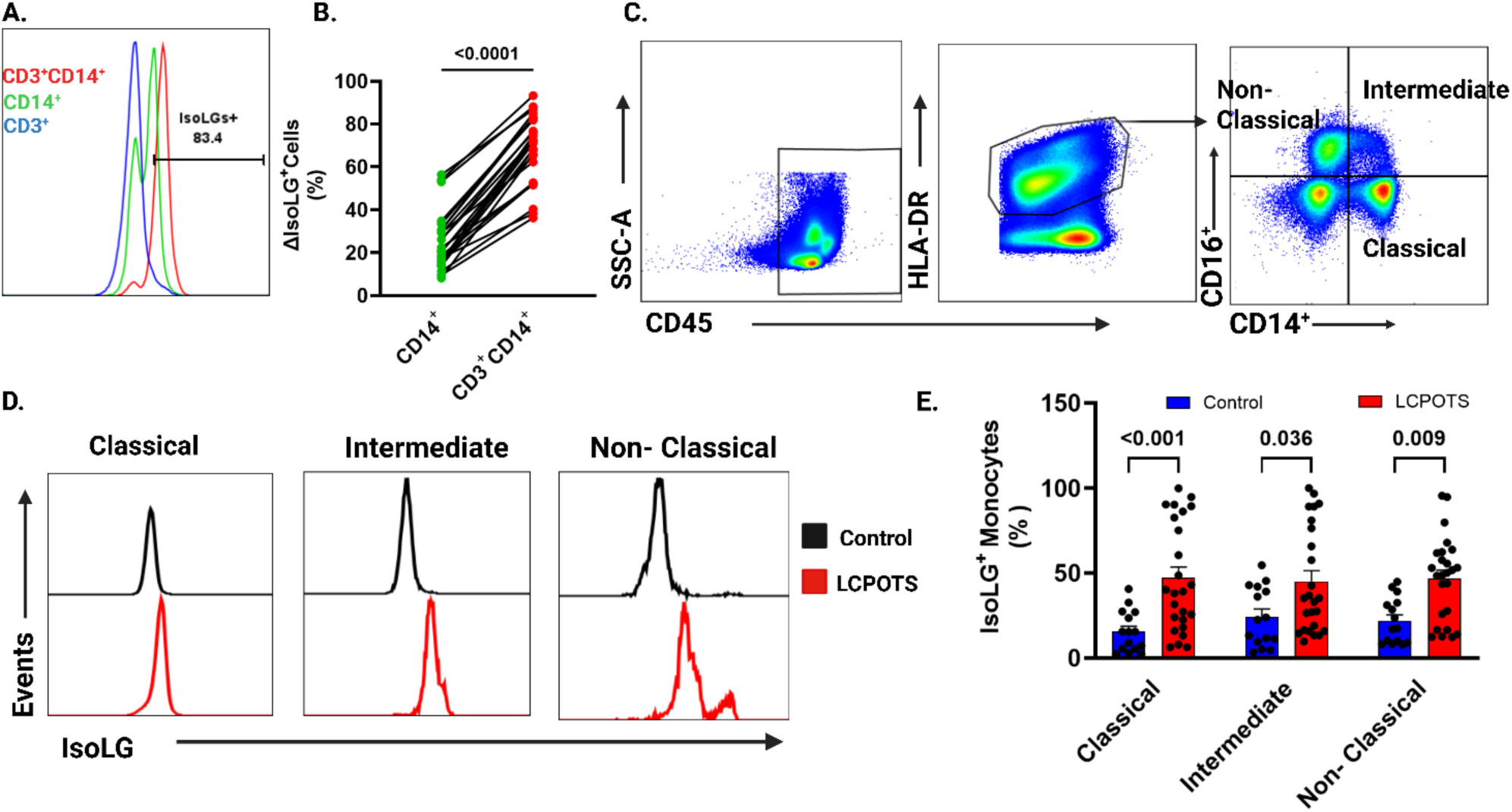
Accumulation of IsoLG-adducts in circulating monocytes from patients with LCPOTS. **(A)** Representative overlaid histograms showing intracellular IsoLG staining in CD3^+^ cells (blue), CD14^+^ monocytes (green), and CD3^+^CD14^+^ doublets (red), with the gate used to define IsoLG^+^ events. **(B)** Paired comparison of the frequency of IsoLG^+^ monocytes in CD14^+^ singlet monocytes versus monocytes within CD3^+^CD14^+^ doublets from matched samples, showing increased IsoLG positivity in doublet-associated monocytes. Individual lines connect matched singlet–doublet pairs from the same participant. **(C)** Representative flow cytometric gating strategy used to identify monocyte subsets, including classical (CD14^++^CD16^-^), intermediate (CD14^++^CD16^+^), and non-classical (CD14^+^CD16^++^) monocytes. **(D)** Representative histograms of intracellular IsoLG staining in classical, intermediate, and non-classical monocytes from control (black) and LCPOTS (red) participants. **(E)** Quantification of the percentage of IsoLG^+^ monocytes within the classical, intermediate, and non-classical monocyte subsets in controls and LCPOTS participants. Bars represent mean ± SEM; individual symbols represent individual participants. For panel **(B)**, paired comparison was performed using a two-sided Wilcoxon signed-rank test. For panel **(E)**, comparisons between groups across the three monocyte subsets were performed using two-way ANOVA followed by Tukey’s multiple-comparisons correction; adjusted *P* values are shown above each comparison. Controls *n* = 15; LCPOTS *n* = 25.

### Plasma and Cellular Cytokine Profiles

Using mesoscale analysis, we found that LCPOTS subjects have a significant increase in multiple circulating pro-inflammatory cytokines compared to controls. Conversely, there were no significant differences between groups for IL-4 or IL-10. These results demonstrate a pro-inflammatory cytokine profile in LCPOTS patients, while anti-inflammatory mediators remain unchanged (Table 2).

In additional studies, we cultured isolated CD4^+^ and CD8^+^ T cells with anti-CD3 and anti-CD28 coated plates for 48 hours and quantified the release of IFN-γ, TNF-α, and IL-17A. (Figure 2S). CD4^+^ T cells from LCPOTS patients produced significantly more pro-inflammatory cytokines than controls (Figure 2S, A). Secretion of IFN-γ and IL-17A was elevated in LCPOTS; TNF-α was not significantly different between groups. A similar pattern was observed for CD8^+^ cells (Figure 2S, B). LCPOTS subjects exhibited significantly elevated IFN-γ and TNF-α, with a trend towards increased IL-17A. Conversely, both CD4^+^ and CD8^+^ T cells from LCPOTS patients showed significantly decreased production of the anti-inflammatory cytokine IL-10 compared to controls (Figure 2S, A-B). These results reveal a pattern of T cell cytokine production in LCPOTS patients that is skewed toward pro-inflammatory and away from anti-inflammatory responses. Responder-strip analysis demonstrated cytokine-specific redistribution at the subject level across tertiles (Figure 2S, C–D). In the CD4^+^ compartment, LCPOTS subjects were enriched in the high tertile for IFN-γ and IL-17A (47% for each cytokine vs 10% in controls) and in the low tertile for IL-10 (50% vs 9% in controls). In the CD8^+^ compartment, LCPOTS subjects were similarly enriched in the high tertile for IFN-γ, TNF-α, and IL-17A, together with a shift of IL-10 toward the low tertile.

### Monocyte Subset Distribution

A comparative analysis of classical (CD14^++^CD16^-^), intermediate (CD14^++^CD16^+^), and non-classical (CD14^+^CD16^++^) showed these were differently distributed between LCPOTS patients and controls (Figure 3S). Non-classical monocytes were significantly higher in LCPOTS patients, while the proportions of classical and intermediate subsets were not different. This shift toward more non-classical monocytes suggests prolonged survival of these cells, as they evolve from classical monocytes (26).

### Transcutaneous vagal nerve stimulation (t-VNS) reduces CD3^+^CD14^+^ doublets, monocyte oxidative stress and inflammatory markers

As shown in Figure 2, subjects with LCPOTS have reduced cardiovagal tone, with little or no change in sympathetic outflow. To determine whether enhancing vagal nerve signaling could reverse the immune phenotype identified in LCPOTS, we treated a subset of our subjects with transcutaneous vagal nerve stimulation (t-VNS) applied to the ear tragus twice daily for 30 minutes for 28 days. Following t-VNS, orthostatic tachycardia during upright HUT was reduced overall after treatment (Figure 7A). The symptom burden, reflected by the Total COMPASS-31 score, improved in most patients after t-VNS (Figure 7B). The frequency of circulating CD3^+^CD14^+^ doublets, expressed as a percentage of live CD45^+^ cells, was reduced compared with baseline (Figure 7C and D). In parallel, the proportion of IsoLG^+^ and IL-6^+^ monocytes, expressed as a percentage of CD14^+^ monocytes, was markedly decreased after t-VNS (Figure 7E and F). Mitochondrial mass, mitochondrial superoxide production, and total cellular ROS were also reduced by t-VNS (Figure 7G–I). We also examined the effect of t-VNS on NRF2-dependent genes and mitochondrial genes using single-cell sequencing in two subjects with LCPOTS before and after t-VNS. In these subjects, t-VNS dramatically reduced mRNAs for mitochondrial proteins and upregulated NRF2-dependent genes (Figure 5S).

**Figure 7:**
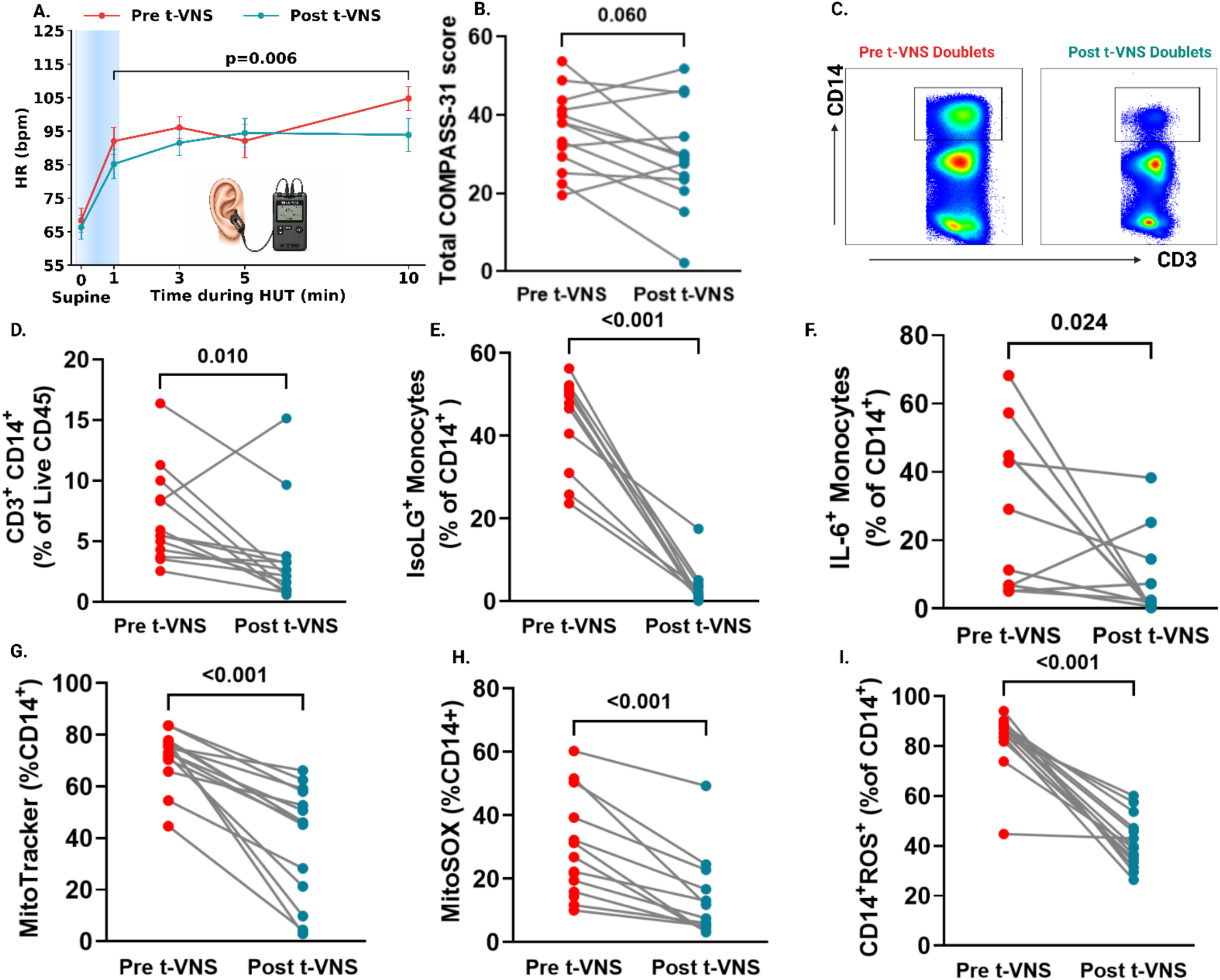
Transcutaneous vagal nerve stimulation (t-VNS) is associated with reduced orthostatic tachycardia, decreased circulating CD3^+^CD14^+^ doublets, and reduced monocyte inflammatory and oxidative stress markers in LCPOTS. **(A)** Heart rate (HR) during head-up tilt (HUT) before and after t-VNS in participants with LCPOTS. Values are shown as mean ± SEM. The shaded region denotes the supine baseline period, a linear mixed-effects model with visit (pre vs post t-VNS, n=12 LCPOTS patients). **(B)** Total COMPASS-31 score before and after t-VNS. **(C)** Representative flow cytometry plots showing the CD3^+^CD14^+^ doublet population before and after t-VNS, using identical gating boundaries. **(D–I)** Paired analyses of CD3^+^CD14^+^ doublets (expressed as a percentage of live CD45^+^ cells) (**D**), IsoLG^+^ monocytes (**E**), IL-6^+^ monocytes (**F**), MitoTracker^+^ monocytes (**G**), MitoSOX^+^ monocytes (**H**), and total ROS^+^ monocytes (**I**) are expressed as a percentage of CD14^+^ monocytes. For **B** and **D–I**, each line represents one participant; paired comparisons were performed using 2-sided Wilcoxon signed-rank tests (n=13 LCPOTS patients). LCPOTS, postural orthostatic tachycardia syndrome; HUT, head-up tilt; HR, heart rate.

## Discussion

Following the SARS-CoV-2 pandemic, there has been a marked increase in the prevalence of POTS. In the present study, we found that these subjects exhibit a state of continued immune activation, characterized by increased circulating cytokines, persistent circulating T cell-monocyte doublets, and a striking increase in the content of IsoLG-adducts within monocytes complexed with T cells in those doublets. Importantly, T cells in these complexes exhibit TH1 and TH17 phenotypes, and the levels of their corresponding cytokines correlate strongly with postural changes in heart rate. These immune abnormalities occurred in parallel with a distinct autonomic phenotype characterized by exaggerated orthostatic tachycardia, greater orthostatic symptom burden, and increased COMPASS-31 scores. Our autonomic data confirm that the dominant abnormality in LCPOTS is reduced cardiovagal modulation rather than a uniformly hyperadrenergic phenotype (27). Measures linked to parasympathetic function, including HF _RRI_, RMSSD, SD1, and baroreflex sensitivity during upward sequences (BRS Seq Up), were lower in LCPOTS, with lower total power as well. In contrast, LF _RRI_, LF/HF ratio, blood pressure variability indices, and Valsalva-derived sympathetic measures were not different between-groups. Importantly, t-VNS in a subset of our LCPOTS subjects led to striking improvement in almost all symptomatic and immune abnormalities observed in this condition.

### T Cell-Monocyte Immune Synapses: A Novel Biomarker for LCPOTS

A central finding of our study is the approximately threefold increase in circulating T cell– monocyte doublets in LCPOTS patients compared with controls. Although historically regarded as technical artifacts in flow cytometry, accumulating evidence indicates that these complexes represent biologically meaningful interactions between T cells and monocytes (25). These conjugates display characteristics of true immunological synapses and are more prevalent during immune activation, such as in active tuberculosis, dengue infection, and post-vaccination. We have also previously observed elevated T cell–monocyte doublets in individuals with HIV infection and glucose intolerance (24). In the present study, we demonstrated increased TCR–HLA interactions within CD3^+^CD14^+^ doublets using fluorescence resonance energy transfer (FRET), supporting the presence of authentic immune synapses rather than random cellular aggregates. A likely cause of sustained doublet formation is that monocytes of LCPOTS patients exhibit a striking increase in IsoLG-adducts, which act as neoantigens. Peptides derived from such post-translationally modified proteins are presented in the context of both class I and class II major histocompatibility complexes and can activate subsets of both CD4^+^ and CD8^+^ T cells. Our prior studies have shown that these post-translationally modified self-antigens contribute to hypertension (20, 22), systemic lupus erythematosus (23), and heart failure (21). IsoLG-adducts not only lead to self-antigen formation, but also disrupt critical transcription factors involved in autoimmunity (23), and their accumulation in myeloid cells is associated with a striking increase in production of T cell skewing cytokines, including IL-6, IL-23, and IL-1β (20). Recent studies have elucidated the constraints of MHC structure required for presentation of IsoLG-adducted peptides (22, 28). Thus, the monocytes in LCPOTS contain both the biochemical substrate for neoantigen formation and the cellular machinery required for sustained antigen presentation. Consistent with this, the association constant (K_a_) between T cells and monocytes was significantly higher in LCPOTS patients, indicating enhanced binding affinity and sustained intercellular engagement in this condition.

Additional studies provided insight into the mechanisms responsible for IsoLG-adduct formation in the monocytes of humans with LCPOTS. Our single-cell transcriptomic analysis showed that mitochondrial gene expression was markedly increased in these individuals compared to control subjects. This was accompanied by a striking decline in expression of several critical antioxidant genes, including SOD2, GPX4, PRDX1, and HMOX1. Notably, there was a concomitant decrease in mRNA content of the master transcription factor NRF2, which controls expression of many of these genes. This combination of increased mitochondria, which are major producers of ROS, and decreased antioxidant defenses provides a setting for enhanced oxidative injury and, importantly, IsoLG-adduct formation. Consistent with this, we observed increased total ROS and mitochondrial superoxide production in CD14^+^ monocytes from humans with LCPOTS. Together, these findings suggest that monocytes in LCPOTS are metabolically and immunologically reprogrammed toward a persistent inflammatory phenotype.

The biological relevance of these circulating immune synapses is further supported by the functional phenotype of T cells within doublets. Compared with singlet T cells from the same individuals, doublet-associated T cells exhibited markedly enhanced pro-inflammatory cytokine production. Both CD4^+^ and CD8^+^ T cells within doublets demonstrated approximately threefold higher frequencies of IFN-γ and IL-17A expression relative to their singlet counterparts. Importantly, the frequency of IFN-γ-producing T cells within doublets correlated strongly with the magnitude of orthostatic heart rate increase, as did the proportion of IL-17A-producing cells. These robust associations support the concept that immune activation in LCPOTS is not merely an epiphenomenon but is mechanistically linked to the severity of autonomic dysfunction. Thus, the doublet-associated inflammatory T cell phenotype tracks with a quantitative measure of disease severity, persistent antigenic stimulation, and oxidative myeloid reprogramming in LCPOTS. Importantly, the association between cytokine-positive doublets and clinical severity was not limited to orthostatic tachycardia. The frequencies of both CD3^+^CD14^+^IFN-γ^+^ and CD3^+^CD14^+^IL-17A^+^ doublets also correlated with total COMPASS-31 score, indicating that this inflammatory doublet phenotype tracks with greater autonomic symptom burden as well as physiological orthostatic heart rate responses. This is relevant because COMPASS-31 is a validated quantitative instrument for assessing autonomic symptoms and has been used to demonstrate substantial symptom burden in POTS (29, 30), supporting the concept that persistent immune activation is linked to the broader symptomatic phenotype of LCPOTS.

In keeping with the concept of persistent antigenic stimulation in LCPOTS, we observed a striking predominance of effector memory EM and TEMRA subsets, accounting for approximately 90% of doublet-associated T cells. This contrasts sharply with the composition of circulating T cells in healthy individuals, where naive and central memory subsets typically predominate (31, 32). TEMRA cells represent a highly differentiated subset with potent effector functions, including rapid cytokine production and cytotoxic activity, but limited proliferative capacity (33), in keeping with the profile of cytokines we observed with intracellular staining in the circulation and released from cultured T cells of LCPOTS subjects. Because TEMRA cells are typically expanded in chronic viral infections, autoimmune diseases, and aging-related inflammation (34), their predominance within T cell-monocyte doublets is more consistent with sustained immune activation than with a transient residual response to prior infection. We considered that this could be due to persistent viral proteins, but in preliminary studies (n = 16), we were unable to detect SARS-CoV spike protein mRNA using droplet digital PCR to screen in monocytes of these subjects (data not shown). Although this does not exclude tissue-restricted viral persistence, it makes ongoing systemic monocyte infection a less likely explanation for the circulating immune phenotype observed here.

An important finding in our study is that a one-month treatment with transcutaneous vagal nerve stimulation markedly improved symptoms, reduced orthostatic tachycardia, and reduced many of the immunological parameters observed in subjects with LCPOTS. Vagal stimulation has proven beneficial for multiple inflammatory conditions including rheumatoid arthritis (35), systemic lupus (36), osteoarthritis (37), and other chronic diseases. Importantly, Stavrakis et al found that t-VNS improved postural tachycardia, reduced antiadrenergic autoantibodies and inflammatory cytokines in subjects with POTS (38). While t-VNS likely has several actions, engagement of neural circuits that reduce inflammation, as proposed by Tracey and colleagues, likely plays a critical role (39). These studies have shown that immune/cholinergic interfaces with myeloid cells modulate their inflammatory phenotype. Compatible with our study, Han et al showed that α7 nicotinic acetylcholine receptor signaling increased antioxidant genes and reduced cytokine production by macrophages in an ischemic stroke model (40).

Our data are compatible with a paradigm in which reduced vagal tone, through mechanisms yet to be defined, leads to mitochondrial expansion in monocytes, increased mitochondrial ROS production, IsoLG formation, neoantigen formation, and persistent T cell activation. The vagal nerve exerts well-described anti-inflammatory effects on myeloid cells (41), and thus impaired cardiovagal modulation can remove this important restraint on monocyte activation, cytokine production, and T cell–monocyte interaction. It is also possible that the increase in circulating and immune cells-derived cytokines further impairs cardiovagal control in a feed-forward fashion.

Both IFN-γ and IL-17A have been implicated in neuroinflammation and central nervous system demyelination in various autoimmune and post-infectious conditions (16, 42–44). These promote blood-brain barrier disruption, microglial activation, and recruitment of peripheral immune cells into the central nervous system, potentially affecting autonomic centers in the brainstem and hypothalamus that regulate cardiovascular function. Inflammatory cytokines can also directly affect peripheral autonomic neurons and ganglia. IFN-γ has been shown to modulate sympathetic neurotransmission and alter norepinephrine release, while IL-17A can induce peripheral nerve inflammation and damage in animal models (45, 46). In this framework, impaired cardiovagal function and immune activation may reinforce one another in a feed-forward manner.

Many of our findings in humans with LCPOTS resemble features of myalgic encephalomyelitis/chronic fatigue syndrome (ME/CFS), which is characterized by profound fatigue, autonomic dysfunction, and immune dysregulation (47, 48). The mechanistic overlap between LCPOTS and ME/CFS, including oxidative stress, mitochondrial dysfunction, and T cell activation, suggests shared pathogenic pathways that may respond to similar therapeutic approaches. Notably, a subset of LCPOTS patients meets diagnostic criteria for both POTS and ME/CFS, further supporting mechanistic convergence between these syndromes (47). Recently, Shankar *et al.* have shown that T cells from subjects with Long COVID and subjects with ME/CFS exhibit increased ROS production, lipid peroxidation, and evidence of mitochondrial dysfunction in human lymphocytes with either condition (19). We speculate that the combination of increased mitochondrial biogenesis and uncoupled, superoxide-generating respiration in circulating monocytes serves a dual pathogenic role in LCPOTS: generating IsoLG-protein adducts that act as neoantigens to drive pathogenic T cell responses, while simultaneously representing a wasteful, inefficient use of systemic energy reserves; a mechanism that, given the outsized (∼25%) contribution of immune cells to whole-body energy expenditure, could directly contribute to the profound fatigue these patients experience.

This study complements and extends prior work implicating immune dysregulation and autoimmunity in Long COVID-associated autonomic dysfunction and POTS. Reviews of Long COVID and POTS have highlighted generalized immune dysregulation and the presence of multiple autoantibodies, and recent case-control data have identified circulating autoantibodies against vasoactive targets relevant to orthostatic intolerance (49, 50). Similarly, Wallukat *et al.* identified functional autoantibodies against adrenergic and muscarinic receptors in Long COVID patients with autonomic symptoms (51). We have also shown that antibodies to IsoLG-adducts are formed in inflammatory diseases (23), and it is interesting to speculate that epitope spreading resulting from B cell uptake of IsoLG-adducted proteins could initiate the formation of such autoantibodies to autonomic receptors. In accord with this, Guedes de Sá recently showed that subjects with long COVID have a gamut of autoantibodies against central nervous system and peripheral nervous system proteins (52).

Peripheral and autonomic small-fiber involvement has also been documented in Long COVID, including cohorts with dysautonomic symptoms and histologic evidence of autonomic small-fiber loss (53, 54). The elevated pro-inflammatory cytokines, particularly IL-17A and IFN-γ, are consistent with mechanisms that promote neuroinflammation, impair Schwann-cell-mediated myelination, and contribute to neuropathic injury, thereby providing a plausible mechanistic link to small-fiber involvement in LCPOTS (46). Although we did not directly assess small-fiber structure or function, the present findings are consistent with an inflammatory milieu capable of promoting peripheral autonomic nerve injury. We propose that impaired cardiovagal regulation stimulates monocyte ROS production, promotes neoantigen formation, and enhances T cell activation. This persistent immune response, together with disrupted mitochondrial function, likely contributes to the multisystem manifestations of LCPOTS. Several limitations of this study should be acknowledged. While we observed strong associations between immune activation and clinical and physiological measures of orthostatic intolerance, causality cannot be established from this cross-sectional study. In addition, we focused on circulating immune cells and plasma mediators and did not assess tissue-level immune infiltration in autonomic ganglia or central nervous system structures, which may represent important sites of immunopathology. Our subjects were taking a variety of medications that might have affected their hemodynamic and immune status; however, we stopped all drugs 2 weeks prior to the study in order to minimize their influence. Several mechanistic analyses were also performed in smaller subsets of the cohort, and complete autonomic profiling was not feasible in all participants because some were unable to complete the protocol or experienced worsening of symptoms during testing. Finally, scRNA-seq was restricted to a small subset and was performed only in women, consistent with the strong female predominance of LCPOTS/POTS.

Together, our findings support a model in which reduced cardiovagal restraint permits a self-sustaining cycle of monocyte mitochondrial expansion, oxidative stress, IsoLG-neoantigen formation, and sustained T cell activation. The expanded and bioenergetically inefficient immune compartment likely imposes a systemic energy cost, offering a plausible physiological mechanism for the profound fatigue that accompanies LCPOTS. The responsiveness of this condition to non-invasive vagal neuromodulation, evidenced by reductions in doublet frequency and IsoLG^+^/IL-6^+^ monocytes following t-VNS, indicates that the vagal–monocyte–T cell circuit described here is not merely correlative but represents a therapeutic target. Larger longitudinal studies, including interventions that scavenge IsoLG-adducts, restore NRF2-dependent antioxidant defenses, or directly engage cardiovagal afferents, will be needed to establish causality and determine whether disrupting this cycle can durably improve the autonomic, immunologic, and fatigue-related burden of this prevalent post-viral syndrome.

## Methods

### Sex as a biological variable

This study enrolled both male and female participants, reflecting the well-documented strong female predominance of POTS and LCPOTS. All LCPOTS patients were female. Because the LCPOTS cohort was exclusively female, formal sex-stratified comparisons between LCPOTS and controls were not possible, and the findings reported here should be interpreted as applicable primarily to female participants. Single-cell RNA sequencing was likewise performed only in a subset of female participants. This sex distribution is consistent with the epidemiology of LCPOTS.

### Study design

This was a case-control study involving 25 LCPOTS subjects and 15 subjects who recovered from COVID without any sequela, enrolled at the Vanderbilt Autonomic Dysfunction Center between 2023 and 2025. LCPOTS was defined as the presence of orthostatic tachycardia (heart rate increase >30 bpm within 10 minutes of standing) persisting for >3 months following RT-PCR-confirmed SARS-CoV-2 infection, without orthostatic hypotension (systolic blood pressure drop >20 mmHg or diastolic drop >10 mmHg). Controls were individuals without symptoms of POTS and who were confirmed not to have orthostatic tachycardia. As with most of the United States population, all control subjects had previously had a SARS-CoV-2 infection and recovered without post-COVID sequelae, confirmed by clinical assessment and absence of Long COVID symptoms for at least 3 months post-infection.

Subjects were asked to abstain from exercise for at least 12 hours before the study. Multivitamins, non-steroidal anti-inflammatory drugs (i.e., aspirin, ibuprofen), alcohol, and caffeine were withheld for at least 72 hours before the hemodynamic measurements. Medications that affect autonomic function, heart rate, blood pressure, and blood volume were discontinued for 5 half-lives before the study. Subjects were studied in the fasting condition.

Autonomic function tests were performed at Vanderbilt Autonomic Dysfunction Center (ADS) following standardized protocols to assess adrenergic and cardiovagal responses (2). Autonomic evaluation included hemodynamic measurements obtained while supine and then at 1, 3, 5, and 10-minute head-up tilt at 75 degrees. While in the supine position, heart rate response to deep breathing (respiratory sinus arrhythmia) and the Valsalva maneuver (VM) were measured to assess parasympathetic activity (55). Brachial blood pressure and heart rate were measured using an automated brachial sphygmomanometer (Dinamap, GE Medical Systems Information Technologies, Milwaukee, WI). A Finapress® device (BMEYE, Amsterdam, the Netherlands) was used to record continuous blood pressure using the finger volume clamp method. Data were collected using the WINDAQ (Akron, USA) data acquisition system. The changes in heart rate (ΔHR) in response to orthostatic changes were used as an index of autonomic function as described by Norcliffe-Kaufmann et al (56). The COMPASS-31 and the Vanderbilt Orthostatic Symptom Score (VOSS) were used to assess autonomic symptom burden during the study visit (29, 57). VOSS was completed before any orthostatic challenge testing to establish baseline orthostatic symptom severity. We excluded individuals with a history of atrioventricular block, myocardial infarction, angina, heart failure, pacemaker, stroke, transient ischemic attack within 6 months before enrollment, history or current seizures, uncontrolled hypertension (BP>140/90 despite appropriate treatment), type 1 or type 2 diabetes mellitus, impaired hepatic function, impaired renal function test (eGFR<60 mL/min/1.73m2), anemia (hemoglobin <10 g/dl), use of immunosuppressive medications, central acetylcholinesterase inhibitors, pregnant or breastfeeding women and autoimmunity, or inability to provide informed consent.

### Transcutaneous vagal nerve stimulation (t-VNS)

In a subset of participants with LCPOTS, t-VNS was delivered using an FDA-approved TENS 7000 device fitted with ear-clip electrodes applied to the auricular tragus. Stimulation was delivered at 30 Hz with a pulse width of 300 µs, and the amplitude was adjusted to each participant’s perception threshold. Participants underwent 30 minutes of stimulation twice daily for 28 days. Symptoms and the inflammatory/immune phenotype were assessed before and after the intervention.

### Heart Rate Variability and Valsalva Maneuver Analysis

Continuous electrocardiogram (ECG; Ivy 450C, Ivy Biomedical Systems, Inc., Branford, CT, USA) and finger blood pressure (FBP; BMEYE, Amsterdam, The Netherlands) were recorded at a sampling frequency of 1 kHz using WINDAQ software (DATAQ Instruments, Akron, OH, USA). Intermittent brachial blood pressure was measured on the contralateral arm (Ivy 450C) before each autonomic function test to cross-calibrate the finger blood pressure signal. Data were analyzed using Physiowave©, a custom MATLAB-based software (A. Diedrich, Vanderbilt University Medical Center, TN, USA) (58). R-waves, systolic blood pressure (SBP), and diastolic blood pressure (DBP) were automatically detected from the waveforms and visually inspected for artifacts. Beat-to-beat time series were validated, interpolated, and resampled at 5 Hz. Baseline parameters were calculated over a 5-minute resting period. Power spectral density of R-R intervals and blood pressure was estimated using Welch’s Fast Fourier Transform (FFT) method (59). Spectral power was integrated for very low frequency (VLF: 0.003–0.04 Hz), low frequency (LF: 0.04–0.15 Hz), and high frequency (HF: 0.15–0.40 Hz) bands according to established guidelines (60).

Spontaneous BRS was assessed using the sequence method, defined as at least three consecutive heartbeats where SBP and the subsequent R-R interval changed in the same direction (thresholds: >1 mmHg and >1 ms, respectively). Slopes with a correlation coefficient (r>0.85) were averaged to determine BRS during rising pressure (BRSup) and falling pressure (BRSdown) (61). Additionally, the BRS alpha index was calculated as the square root of the ratio of R-R interval power to SBP power within the LF and HF bands (62).

For the Valsalva maneuver (VM) analysis, the following distinct phases were defined: a baseline period (45 to 15 s prior to straining), Phase 1 (VM1; onset of strain), and Phase 2 (VM2; from maximum BP to the end of strain). Phase 2 was further subdivided into early (VM2e) and late (VM2l) phases at the SBP nadir. Phase 3 (VM3) was defined from the release of strain to the point of minimum BP, while Phase 4 (VM4) was the period of BP recovery, overshoot, then return to baseline (63).

### Single-cell RNA-sequencing

Single-cell RNA sequencing was performed on PBMCs from 4 LCPOTS and 4 control subjects. Single cells from each participant were barcoded using TotalSeq-C hashtag antibodies (BioLegend) to enable sample multiplexing and batch effect correction.

Single-cell sequencing libraries were prepared using the Chromium Single-Cell 5’ v2 Chemistry Library and Gel Bead Kit (10x Genomics) following the manufacturer’s protocol. 30,000 PBMCs were loaded per well to ensure comprehensive immune cell profiling while maintaining optimal cell recovery (64). Libraries were sequenced on the NovaSeq 6000 platform (Illumina) with a target sequencing depth of 50,000 reads per cell (65).

Raw BCL files were demultiplexed and aligned to the human reference genome (GRCh38) using Cell Ranger Single Cell Software v6.0.0 (10x Genomics). Quality control, data normalization, dimensionality reduction, and differential gene expression analysis were performed using Seurat v5.3 in R v4.2.0 (66). Cells were filtered to include only those with detected RNA features between 200 and 5,000 genes, total UMI counts between 1,000 and 50,000, and mitochondrial gene content <10% to exclude low-quality cells. Data normalization was performed using the Log Normalize method, and the top 2,000 highly variable features were identified using the Find Variable Features function. Principal component analysis was conducted using the Run PCA function, and the first 30 principal components were used for downstream analysis. An elbow plot was used to guide cluster segregation (Figure 4S-A). Dimensionality was set at 1:10, and cell clustering was performed using the Find Neighbors and Find Clusters functions with a resolution of 0.1. Clusters were visualized using Uniform Manifold Approximation and Projection (UMAP) and were identified using canonical markers. Differential gene expression of monocytes was compared using the Wilcoxon rank-sum test and visualized using a volcano plot. Gene Ontology and KEGG analyses were performed using ShinyGO 0.85.

### Oxidative Stress and Mitochondrial Staining in CD14^+^ Monocytes

PBMCs were isolated as described above. Following viability and surface-marker staining, cells were used to measure total intracellular reactive oxygen species (ROS), mitochondrial superoxide, or mitochondrial content by flow cytometry. In all assays, CD14^+^ monocytes were identified based on surface staining and analyzed as live, unfixed cells.

### Total Cellular ROS Detection

Total intracellular reactive oxygen species in CD14^+^ monocytes were assessed using the Total Reactive Oxygen Species (ROS) Assay Kit 520 nm (Invitrogen, catalog #88-5930-74) by flow cytometry. After viability and surface staining, cells were resuspended in ROS Assay Buffer. The ROS Assay Stain Concentrate was reconstituted in freshly opened dimethyl sulfoxide (DMSO) to prepare a 500X stock solution. Cells were incubated with 1X ROS Assay Stain (100 µL per sample) for 60 minutes at 37°C with 5% CO_2_, protected from light. Samples were then analyzed immediately by flow cytometry using the FITC channel.

### Mitochondrial Superoxide Detection (MitoSOX)

Mitochondrial superoxide production in CD14^+^ monocytes was measured using MitoSOX™ Red Mitochondrial Superoxide Indicator (Invitrogen, catalog #M36008) by flow cytometry. MitoSOX was reconstituted in DMSO to prepare a stock solution(5mM) used to prepare a (1 µM) working solution in HBSS. After viability and surface staining, cells were incubated with MitoSOX (1 ml/sample) at 37°C with 5% CO_2_ for 20 minutes, protected from light. Following incubation, cells were washed three times with cold MACS buffer to remove unbound dye and immediately resuspended for analysis. All measurements were performed on live, unfixed cells. CD14^+^ monocytes were identified by surface staining and gated for analysis.

### Fluorescent labeling of mitochondria in live cells

Mitochondrial content in CD14^+^ monocytes was assessed using MitoTracker™ Green FM (Thermo Fisher Scientific, catalog # M7514), which was reconstituted in DMSO according to the manufacturer’s instructions to generate a 1 mM stock solution. For mitochondrial staining, a final 5 nM staining solution was prepared fresh in HBSS. Cells were used after viability and surface staining, resuspended in prewarmed (37°C) staining solution (1ml/sample), and incubated for 15–45 min under standard culture conditions. Cells were then pelleted, resuspended in MACS buffer, and analyzed by flow cytometry.

### Flow cytometry

Flow cytometry was performed on PBMCs as previously described using the fluorophores shown in Table 1s. Blood samples were obtained from all participants, and peripheral mononuclear cells (PBMCs) were separated using standard Ficoll-Paque density gradient centrifugation. Dead cells were excluded, and gates were established by Fluorescence minus one (FMO) controls. Data were acquired on a Cytek Aurora flow cytometer and analyzed using FlowJo software. T cell-monocyte doublets were identified as events that stained positively for the pan T cell marker CD3 and the pan monocyte marker CD14 and expressed as a percentage of total live leukocytes (CD45). To quantify the strength of association between T cells and monocytes, we calculated the T cell-monocyte association constant (T:M K_a_) as previously described (25). T:M K_a_ was defined as the frequency of CD3^+^CD14^+^ complexes divided by the product of the frequencies of CD3^+^CD14^−^ T cells and CD3^−^CD14^+^ monocytes. Frequencies were calculated from live CD45^+^ events. We employed a flow cytometry-based Förster resonance energy transfer approach between anti-pan class I HLA and anti-TCRα/β to estimate true immunological synapse formation between monocytes and T cells. Full list of antibodies used in Table 1S. Intracellular IsoLG-adducted proteins were quantified by flow cytometry using the D11 single-chain antibody (ScFv) as previously described (20). D11 recognizes IsoLG-lysine adducts independent of adjacent amino acid sequence or protein backbone. After surface-marker staining, cells were fixed, permeabilized, and incubated with fluorochrome-conjugated D11. IsoLG positivity was quantified within CD14^+^ monocytes and the indicated monocyte subsets according to labeled D11 FMO control.

For the paired t-VNS analysis, pre- and post-intervention samples were processed using identical staining panels and gating strategy. Circulating CD3^+^CD14^+^ doublets were quantified as a percentage of live CD45^+^ cells, whereas IsoLG^+^ and IL-6^+^ monocytes were quantified as percentages of CD14^+^ monocytes.

### Cytokine measurement

Plasma cytokines were measured using the Meso Scale Discovery (MSD-ECL, Rockville, MD) and the U-PLEX Custom Biomarker (hu) kit according to manufacturer instructions. We quantified circulating levels of IFN-γ, IL-1β, IL-4, IL-6, IL-8, IL-10, IL-12/IL-23p40, IL-17A, and TNF-α using this approach. To determine whether LCPOTS is accompanied by functional reprogramming of the adaptive immune compartment, CD4^+^ and CD8^+^ T cells were isolated from human PBMCs by negative selection using the CD4^+^ T Cell Isolation Kit, human (Miltenyi Biotec, 130-096-533) and the CD8^+^ T Cell Isolation Kit, human (Miltenyi Biotec, 130-096-495), activated *ex vivo* with anti-CD3/anti-CD28, and cultured separately for 72 hours. A four-cytokine panel (IFN-γ, TNF-α, IL-17A, IL-10) was quantified in culture supernatants using Meso Scale Discovery as previously mentioned.

### Statistical Analyses

Statistical analyses were performed using GraphPad Prism version 10.0 (GraphPad Software, San Diego, CA), R version 4.2.0 (R Foundation for Statistical Computing, Vienna, Austria), and Python version 3.12 with the statsmodels (v0.14.6) library. Data are presented as mean ± standard deviation (SD) for demographic characteristics and as mean ± standard error of the mean (SEM) for hemodynamic repeated-measures data. Other experimental data are presented as median [Q1, Q3] or mean ± SEM, as indicated. For box-and-whisker plots, boxes represent the 25th–75th percentiles, center lines represent the median, whiskers extend to 1.5 × IQR, and individual points represent participants. For HRV and Valsalva maneuver analyses, prespecified quality-control procedures were applied before statistical testing, including handling of extreme values according to the 1.5 × IQR rule. Repeated-measures analysis of hemodynamic data during head-up tilt (HUT; heart rate, systolic blood pressure, and diastolic blood pressure at 1, 3, 5, and 10 min) was performed using a Gaussian generalized estimating equation (GEE) model with exchangeable working correlation structure and robust standard errors. Reported *P* values represent the overall between-group effect across upright time points. Two-group comparisons for continuous variables, flow cytometry outcomes, and the T cell–monocyte association constant were performed using two-sided Mann–Whitney U tests. Paired within-subject comparisons were analyzed using two-sided Wilcoxon signed-rank tests. Comparisons across multiple related groups were analyzed using the Friedman test with Dunn’s multiple-comparisons correction. Monocyte subset analyses were performed using two-sided exact Mann-Whitney U tests for each subset, while comparisons of IsoLG positivity across monocyte subsets and groups were performed using two-way ANOVA with Tukey’s multiple-comparisons correction. Associations between continuous variables were assessed using Spearman’s rank correlation. scRNA-seq differential-expression analyses were performed using the Wilcoxon rank-sum test with Bonferroni correction, and GO/KEGG enrichment analyses were corrected using the Benjamini–Hochberg false-discovery rate. Cytokine concentrations were analyzed using distribution-free methods and were also presented as subject-level frequency distributions. Between-group differences in each cytokine were tested using two-sided exact Mann-Whitney U tests; p-values are reported. For *ex vivo*–cultured T cells, cytokine levels in the supernatant were also presented as subject-level frequency distributions to visualize between-group shifts in T cell phenotype. Each cytokine was additionally expressed as a categorical variable, a strategy widely used for inflammatory biomarkers in cardiovascular studies (67, 68), in which the pooled Control and LCPOTS values were ranked and partitioned at the 33rd and 67th percentiles, generating three equal-frequency strata: Low (T1; ≤ 33rd percentile), Intermediate (T2; > 33rd to ≤ 67th percentile), and High (T3; > 67th percentile). The proportion of subjects in each tertile was then computed separately for Control and LCPOTS, and the results were displayed as stacked responder-strip plots. Between-group comparisons were performed on the underlying continuous concentration values, not on the tertile categories. For the paired t-VNS analyses, within-subject changes in circulating CD3^+^CD14^+^ doublets, IsoLG^+^ monocytes, and IL-6^+^ monocytes were assessed using two-sided Wilcoxon signed-rank tests.

### Study approval

The study was approved by the Vanderbilt University Institutional Review Board for Human Research. All participants provided written informed consent. The protocol was part of a registered clinical trial (**NCT05421208**).

## Data Availability

All data will be made available upon reasonable request.

https://www.ncbi.nlm.nih.gov/biosample/60208392

## Data availability

Raw single-cell RNA sequencing data generated in this study have been deposited in NCBI Sequence Read Archive (SRA) under BioProject accession PRJNA1464136 (BioSamples SAMN60208390–SAMN60208397), comprising 4 control and 4 LCPOTS participants. Values for all data points in graphs are reported in the Supporting Data Values file.

## Author contributions

MAA designed and performed the experiments, analyzed and interpreted the data, generated the figures, and wrote the manuscript. MV coordinated the clinical study, including participant recruitment, scheduling, and regulatory compliance. YAV performed venipuncture and processed blood samples for downstream analyses. SYP performed head-up tilt testing and additional autonomic function testing. AD and SK acquired, processed, and analyzed heart rate variability and baroreflex data. CNW and SD, provided scientific consultation, methodological guidance, and critical input on study design and interpretation. KCH, PEM, and TXS provided technical and experimental assistance. CAS and DGH supervised the project, secured funding, performed research and data interpretation and contributed to writing the manuscript. All authors critically reviewed the manuscript, contributed intellectual content, and approved the final version for submission.

## Acknowledgments.

Single-cell RNA sequencing was conducted in Vanderbilt Technologies for Advanced Genomics (VANTAGE) at Vanderbilt University Medical Center

