## Supplemental files for "Monocyte Oxidative Stress Underlies Persistent Immune Activation in Long COVID Postural Orthostatic Tachycardia Syndrome"

From the [1]Division of Genetic Medicine & Clinical Pharmacology, Department of Medicine, Vanderbilt University Medical Center, Nashville, TN, 37232, [2]Department of Pharmacology and Biochemistry, Faculty of Pharmacy, Horus University-Egypt, New Damietta 34518, Egypt, [3] Division of Infectious Diseases, Vanderbilt University Medical Center, Nashville, Tennessee, [4]The Center for AIDS Health Disparities Research, Nashville, Tennessee, [5] Veteran's Health Administration, Tennessee Valley Healthcare System, Nashville, Tennessee

Address for Correspondence:

David G. Harrison

Director, Division of Clinical Pharmacology

Room 536 Robinson Research Building

Vanderbilt University Medical Center

Nashville, TN 37232-6602

**Table 1S: Fluorophores used for flow cytometry**

**SURFACE STAINING**

| | **Marker** | **Fluorophore** | **Clone** | **Catalog number** | **Company** | **Staining Concentration** |
|---|---|---|---|---|---|---|
| **1** | Live/Dead Fixable Viability Kit | LIVE/DEAD™ Fixable Violet Dead Cell Stain Kit | N/A | L34955 | Invitrogen | 1:1000 uL of Staining Buffer |
| **2** | Anti-human Anti-CD45 | BV510 Pacific Orange | 2D1 HI30 | 368526 MHCD4530 | Biolegend ThermoFisher Scientific | 1:100 uL of Staining Buffer |
| **3** | Anti-human Anti-CD14 | APC/Cy7 AF700 | QA18A22 6103 | 398708 56-0149-42 | Biolegend InvitrogenPE | 1:100 uL of Staining Buffer |
| **4** | Anti-human Anti-CD16 | BV605 | 3G8 | 302040 | Biolegend | 1:100 uL of Staining Buffer |
| **5** | Anti-human Anti-HLA-DR | PE/Cy7 | 1243 | 307616 | Biolegend | 1:100 uL of Staining Buffer |
| **6** | Anti-human Anti-CD3 | BV786 | UCHT1 | 417-0038-42 | ThermoFisher Scientific | 1:100 uL of Staining Buffer |
| **7** | Anti-human Anti-CD4 | PE | RPA-T4 | 12-0049-42 | Invitrogen | 1:1000 uL of Staining Buffer |
| **8** | Anti-human Anti-CD8 | PerCP/Cy5.5 | SK1 | 344710 | Biolegend | 1:100 uL of Staining Buffer |
| **9** | Anti-human Anti-HLA-A,B,C | APC/Cy7 | W6/32 | 311426 | Biolegend | 1:100 uL of Staining Buffer |
| **10** | Anti-human Anti- TCR α/β | BV 605 | IP26 | 306731 | Biolegend | 1:100 uL of Staining Buffer |
| | **Intracellular Staining** | | | | | |

| | Marker | Flourochrome | Clone | Catalog number | Company | Staining Concentration |
|---|---|---|---|---|---|---|
| **1** | D-11 | Alexa Flour 488 | N/A | N/A | N/A | 1:100 uL of Staining Buffer |
| **2** | IL-6 | APC | MQ2-13A5 | 501112 | Biolegend | 1:100 uL of Staining Buffer |
| **3** | IL-17A | APC | BL168 | 512333 | Biolegend | 1:100 uL of Staining Buffer |
| **4** | IFN-ɣ | Alexa Flour 488 | 4S.B3 | 502517 | Biolegend | 1:100 uL of Staining Buffer |

**Table 2S: Differential expression of antioxidant-related genes in $CD14^{+}$ monocytes from women with LCPOTS versus control subjects**

| Gene symbol | log2 fold change (LCPOTS vs control) | −log10 adjusted P value |
|---|---|---|
| **PRDX1** | **-1.62** | **308.000** |
| **SOD2** | **-1.65** | **308.000** |
| **GPX4** | **-0.78** | **278.209** |
| **SRXN1** | **-2.54** | **253.943** |
| **NFE2L2** | **-0.58** | **91.670** |
| **NQO1** | **-2.22** | **50.677** |
| **HMOX1** | **-0.21** | **21.028** |

LCPOTS: long COVID postural orthostatic tachycardia syndrome. Statistical testing was performed using the Wilcoxon rank-sum test in Seurat with correction for multiple comparisons.

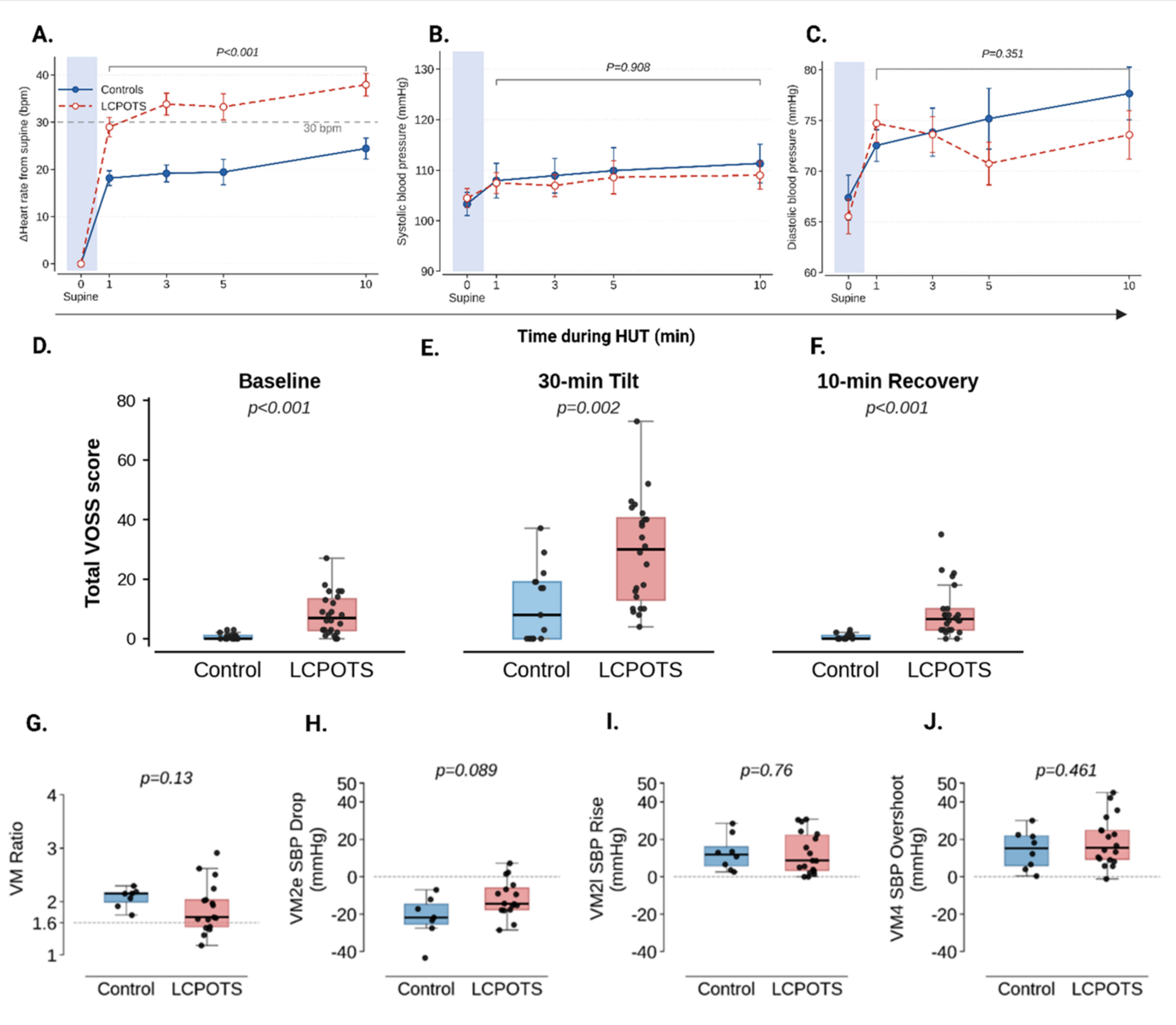

**Figure 1S: Autonomic, Hemodynamic, and Symptom Profiles During Orthostatic Stress and Valsalva Maneuver in Healthy Controls and LCPOTS. (A–C)** Hemodynamic responses during head-up tilt (HUT) in healthy controls (blue) and participants with long COVID postural orthostatic tachycardia syndrome (LCPOTS, red). **(A)** Change in heart rate from supine (ΔHR, bpm) at 1, 3, 5, and 10 minutes of HUT. The dashed horizontal line at 30 bpm indicates the diagnostic threshold for postural tachycardia syndrome. **(B)** Systolic blood pressure (SBP, mmHg) and **(C)** diastolic blood pressure (DBP, mmHg) during HUT. The shaded region denotes the supine baseline period. Symbols indicate mean ± SEM. Upright values were analyzed using a Gaussian generalized estimating equation (GEE) model with exchangeable correlation structure, adjusted for time; the displayed *p* values represent the overall between-group effect across upright time points (1, 3, 5, and 10 min). **(D–F)** Total Vanderbilt Orthostatic Symptom Score (VOSS) measured at **(D)** baseline, **(E)** 30 minutes of head-up tilt, and **(F)** 10 minutes of recovery in the supine position, in controls and LCPOTS participants. **(G–J)** Valsalva maneuver (VM) indices in healthy controls (blue) and LCPOTS participants (red). **(G)** Valsalva ratio (VM Ratio), calculated as the maximum heart rate during phase II divided by the minimum heart rate during phase IV. **(H)** Maximal systolic blood pressure decrease during early phase II (VM2e SBP drop, mmHg). **(I)** SBP rise during late phase II (VM2l SBP rise, mmHg). **(J)** SBP overshoot during phase IV (VM4 SBP overshoot, mmHg). Dashed horizontal lines indicate zero deviation from baseline. For panels (D–J), boxes indicate the interquartile range (25th–75th percentiles), center lines indicate the median, and whiskers extend to the most extreme values within 1.5 × the interquartile range; individual points represent individual participants. Between-group comparisons in (D–J) were performed using two-sided Mann–Whitney U tests; exact *p* values are shown in each panel. Sample sizes varied across panels and time points according to test completion and the availability of analyzable physiologic and symptom data; overall, $n$=7–15 in controls and 16–25 in LCPOTS

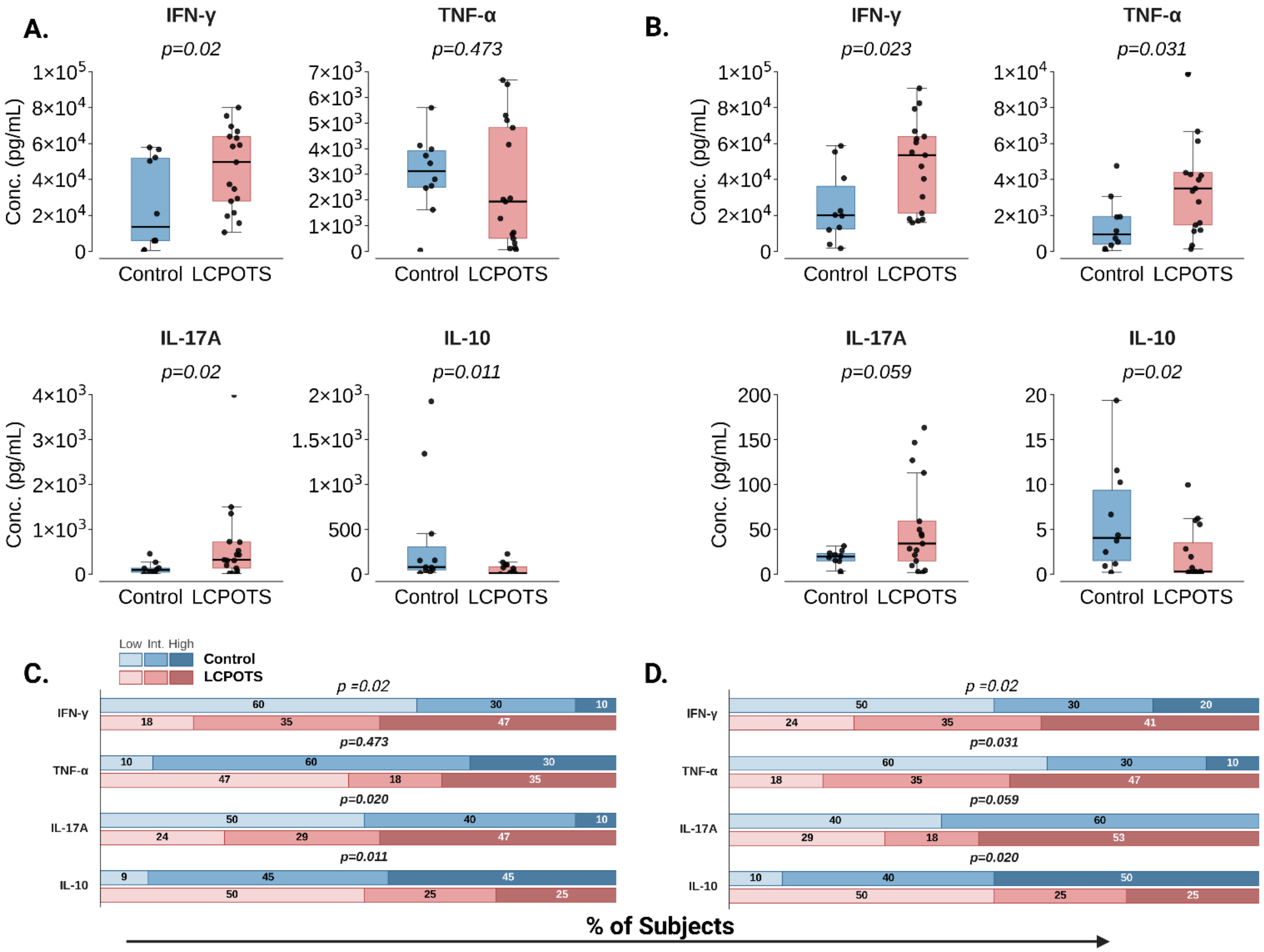

**Figure 2S: Cytokine production by isolated CD4⁺ and CD8⁺ T cells from LCPOTS patients and healthy controls.** Supernatant cytokine concentrations (IFN-γ, TNF-α, IL-17A, IL-10) measured after anti-CD3/anti-CD28 stimulation of isolated CD4+ T cells **(A)** and CD8+ T cells **(B)** from controls (blue) and participants with long COVID postural orthostatic tachycardia syndrome (LCPOTS, red). **(A, B)** Box-and-whisker plots of cytokine concentrations (pg/mL). Boxes indicate the interquartile range (25th–75th percentiles), center lines indicate the median, and whiskers extend to the most extreme values within 1.5 × the interquartile range; individual points represent individual participants. Y-axis scales differ between CD4+ and CD8+ panels to accommodate lower secretion of IL-17A and IL-10 by CD8+ T cells. **(C, D)** Subjectlevel distributions across data-driven tertiles. For each cytokine, all participants (controls + LCPOTS pooled) were ranked by concentration and divided into tertiles at the 33rd and 67th percentiles, defining Low, Intermediate, and High bins. Stacked bars show the percentage of subjects within each tertile for controls (blue, top bar) and LCPOTS (red, bottom bar); color intensity encodes tertile rank (light → dark = Low → High); rows sum to ~100%. Between-group comparisons were performed using two-sided Mann–Whitney U tests on the underlying continuous cytokine values; *p* values are shown above each comparison. Sample sizes: controls *n* = 10–11; LCPOTS *n* = 16–17 (variation reflects cytokine-specific data availability).

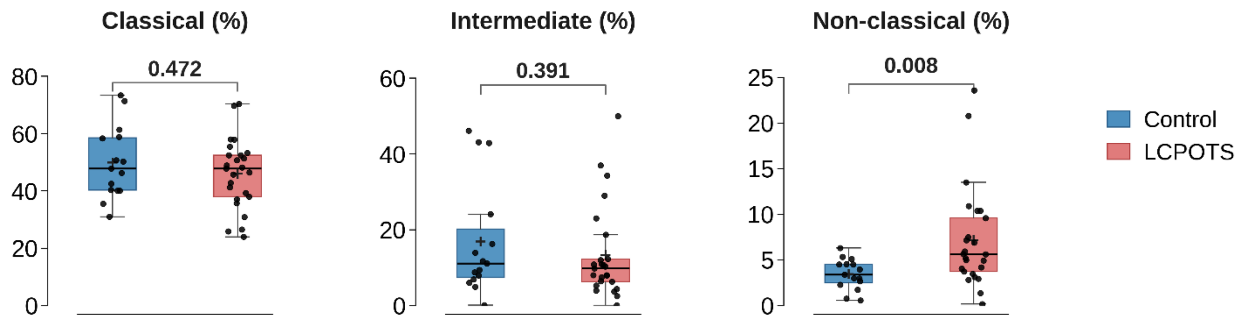

**Figure 3S: Distribution of circulating monocyte subsets in Control and LCPOTS participants.** Relative percentages of classical ($CD14^{++}CD16^{-}$) (left), intermediate ($CD14^{++}CD16^{+}$), and non-classical ($CD14^{+}CD16^{++}$) monocytes (right) within the total circulating monocyte population in controls (blue) and participants with long COVID postural orthostatic tachycardia syndrome (LCPOTS, red). Boxes indicate the interquartile range (25th–75th percentiles), center lines indicate the median, and whiskers extend to the most extreme values within 1.5 × the interquartile range; individual points represent individual participants. Between-group comparisons for each monocyte subset were performed using two-sided exact Mann–Whitney U tests; exact *P* values are shown above each panel. Controls *n* = 15; LCPOTS *n* = 25.

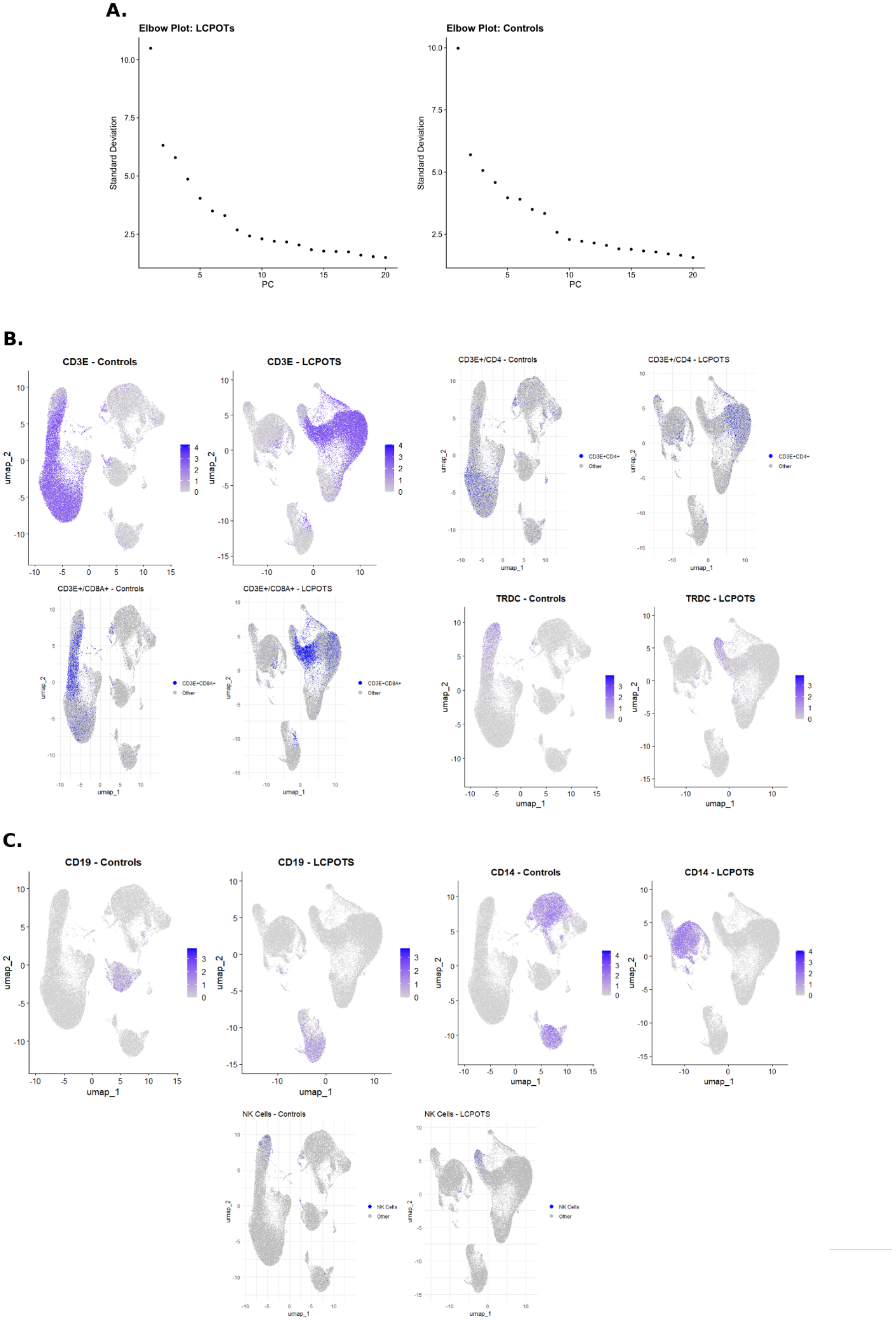
A.
Elbow Plot: LCPOTs
Elbow Plot: Controls
Standard Deviation
PC
B.
CD3E - Controls
CD3E - LCPOTS
CD3E+/CD4 - Controls
CD3E+/CD4 - LCPOTS
CD3E+CD4+
Other
CD3E+/CD8A+ - Controls
CD3E+/CD8A+ - LCPOTS
CD3E+CD8A+
TRDC - Controls
TRDC - LCPOTS
umap_1
umap_2
C.
CD19 - Controls
CD19 - LCPOTS
CD14 - Controls
CD14 - LCPOTS
NK Cells - Controls
NK Cells - LCPOTS
NK Cells

**Figure 4S: Single-cell RNA sequencing (scRNA-seq) clustering and lineage-marker UMAPs in PBMCs from control participants and women with LCPOTS. (A)** Elbow plots showing the standard deviation of the first 20 principal components (PCs) derived from principal component analysis of single-cell transcriptomes, used to identify the inflection point for selecting the number of PCs retained for downstream clustering, in LCPOTS (left) and controls (right). **(B)** UMAP visualizations of T-cell–associated markers and populations, shown separately for controls and LCPOTS. Feature plots display normalized gene-expression intensity (gray → purple gradient) for CD3E (pan-T-cell marker) and TRDC (γδ T-cell T-cell receptor δ constant chain). Highlight plots identify $CD3E^{+}/CD4^{+}$ ($CD4^{+}$ T cells) and $CD3E^{+}/CD8A^{+}$ (CD8+ T cells) populations (purple) relative to all other PBMCs ("Other", gray). **(C)** UMAP visualizations of non-T cell lineage markers and populations, shown separately for controls and LCPOTS. Feature plots display normalized gene-expression intensity (gray → purple gradient) for CD19 (B-cell marker) and CD14 (monocyte marker). Highlight plots identify NK cells (purple) relative to all other PBMCs ("Other", gray). Single-cell RNA sequencing was performed on PBMCs from 4 female control participants and 4 women with LCPOTS. The distribution of major immune lineages, including $CD4^{+}$ and $CD8^{+}$ T cells, γδ T cells, B cells ($CD19^{+}$), monocytes ($CD14^{+}$), and NK cells, was broadly comparable between groups, with no significant differences in overall cell-type proportions. This panel is presented as a descriptive quality-control and cell-annotation figure; formal between-group statistical comparisons of gene expression in $CD14^{+}$ monocytes are presented in Figure 6.

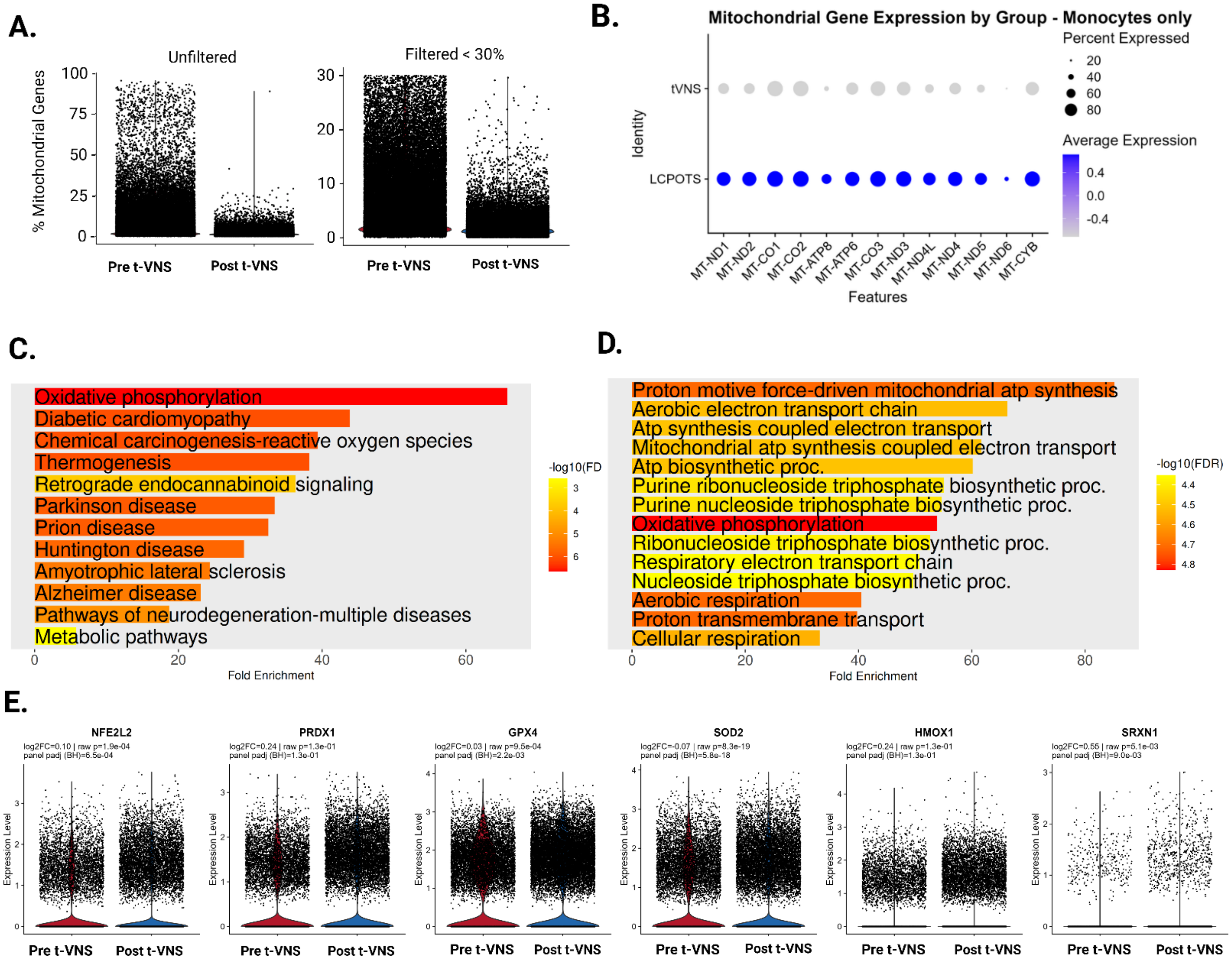

**Figure 5S: t-VNS is associated with reduced mitochondrial transcript burden and altered mitochondrial/oxidative-stress gene signatures in monocytes from LCPOTS.**

**(A)** Percentage of mitochondrial genes detected per cell in monocytes before and after t-VNS, shown in the unfiltered dataset (left) and after filtering cells with mitochondrial gene content <30% (right). **(B)** Dot plot of mitochondrial gene expression in monocytes by group, comparing LCPOTS and t-VNS samples. Dot size indicates the percent of cells expressing each gene, and color indicates average expression. **(C)** KEGG pathway enrichment analysis of differentially expressed mitochondrial-related genes, highlighting enrichment of oxidative phosphorylation and reactive oxygen species–related pathways. **(D)** Gene Ontology biological process enrichment analysis showing overrepresentation of pathways related to mitochondrial ATP synthesis, electron transport, oxidative phosphorylation, aerobic respiration, and cellular respiration. **(E)** Expression of selected oxidative stress– and antioxidant-response genes in monocytes before and after t-VNS, including NFE2L2, PRDX1, GPX4, SOD2, HMOX1, and SRXN1. Exact fold-change and *P* values are shown in each panel.
